# Relationship between calculated total blood count inflammatory readings and the degree of coronary atherosclerosis (Pilot study)

**DOI:** 10.1101/2024.11.02.24316626

**Authors:** Liuizė Agnė, Mongirdienė Aušra, Laukaitienė Jolanta

## Abstract

**Background:** Some calculated total blood count readings are investigated as additional readings to help with evaluation of CAD patients’ clinical management and prognosis.

**Aim:** We aimed to investigate the association between readings such as NLR, MLR, PLR, NMR, LMR, MHR, SII, SIRI and the severity of CAD in patients with SAP.

**Methods:** This retrospective pilot study included 166 patients. All these patients underwent CA or CCTA, or both, to assess severity of CAD. Patients were divided into three ways: 1) according to presence (n=146) or absence (n=20) of CAD; 2) according to Gensini score; 3) according to the CAD-RADS score.

**Results:** Patients with CAD had lower LMR, higher NLR, SIRI, MLR and SII compared to patients without CAD *(p<0.001 and p=0.018,* respectively for SII). According to the CAD severity by Gensini score, the NLR, MLR, SII and SIRI values increases and LMR decreases gradually with severity of CAD (p<0.001). It was found moderate correlation between SII (r=0.511, p*<0.001)*, NLR (r=0.567, p*<0.001)* and SIRI (r=0.474, p*<0.001)* and severity of CAD according to Gensini score. MLR and LMR low correlated with severity of CAD according to Gensini score (r=0.356, p*<0.001*; r=-0.355, p*<0.001,* respectively). The CAD-RADS score weakly correlated with NLR and MHR *(r=0.365, p<0.001; r=0.346, p<0.001*, respectively), and moderately – with LMR, MLR and SIRI (*r=-0.454, p<0.001; r=0.455, p<0.001; r=0.522, p<0.001,* respectively).

**Conclusions:** NLR, LMR and SIRI appear to be potential predictors of chronic inflammation, and SIRI is the best predictor of the degree of atherosclerosis of all the other assessed blood parameters.

## Introduction

Atherosclerosis is a chronic inflammatory process caused by the accumulation of lipids, fibrous elements and calcification in the arteries. This chronic arterial disease starts with endothelial activation, followed by a cascade of events involving vasoconstriction and activation of the inflammatory process leading to the formation of atherosclerotic plaque [1]. Atherosclerosis, as an inflammatory disease, is crucial for the onset and development of coronary artery disease (CAD) because inflammation plays an important role in the formation and progression of atherosclerosis. [2]. CAD remains the leading cause of mortality and morbidity worldwide. Therefore, the identification of high-risk patients with CAD is useful for clinical management and prognosis [3]. Some calculated total blood count readings are investigated as additional readings to help with evaluation of CAD patients’ condition, clinical management, and prognosis.

Markers of inflammatory processes, such as neutrophil-to-lymphocyte ratio (NLR) [27], monocyte-to-lymphocyte ratio (MLR) [24], and platelet-to-lymphocyte ratio (PLR) [25], have been shown to be associated with the severity of CAD and poor cardiovascular prognosis

Studies have shown that immune and inflammatory reactions are closely linked to the development of atherosclerosis [4]. Recently, much attention has been paid to blood cell analysis as a routine laboratory test. Some of the most important cells of the immune system are white blood cells, including lymphocytes, neutrophils, monocytes and macrophages, which play different and important roles in the development of atherosclerosis. For instance, neutrophils can accelerate atherosclerosis at various stages, e.g. by activating macrophages, recruiting monocytes and exerting cytotoxic effects, while lymphocytes modulate the inflammatory response and thus have an anti-atherosclerotic effect [5, 6]. Platelets adhering to the blood vessel wall have been shown to promote leukocyte aggregation and initiate the progression of atherosclerosis before leukocytes penetrate the atherosclerotic plaque [6, 7].

It has been shown that neutrophil, lymphocyte, monocyte and platelet counts may have a prognostic role in the development of CAD, and therefore, as mentioned above, the parameters NLR, MLR and PLR were determined [24, 25, 27]. It has been suggested that they may be more valuable than cell count alone, and many studies have been carried out to assess their evidence as a marker of subclinical inflammation. NLR, PLR were thought to be independent predictors of atherosclerosis severity, and high MLR values can help identify vulnerable plaques in patients with stable angina [3].

A growing number of experimental and clinical investigators confirm that the lymphocyte-to-monocyte ratio (LMR) plays a crucial role in chronic inflammation. Monocytes are pro-inflammatory and differentiate into macrophages in the event of endothelial dysfunction, which subsequently phagocytose lipids in the sub-endothelial space and may differentiate into mast cells and induce atherosclerotic plaque development [21]. Lymphocytes are thought to exert anti-inflammatory effects and regulate the inflammatory response in the pathogenesis of atherosclerosis by enhancing the immune response and are affected by serum catecholamine and cortisol levels during the systemic stress response [29]. Thus, LMR is involved in all stages of coronary atherosclerosis, from initial endothelial dysfunction and plaque disruption to acute atherothrombosis [8].

The neutrophil-to-monocyte ratio (NMR) is also found higher in patients with CAD compared to healthy individuals. This ratio is a marker of systemic inflammation and both neutrophils and monocytes play an important role in the development and progression of atherosclerosis, the main cause of CAD. Elevated NMR is even associated with CAD severity and can therefore be used as a prognostic indicator in these patients. In addition, NMR is generally lower in healthy people, suggesting a more balanced inflammatory response [9].

Monocyte-to-high-density lipoprotein cholesterol (HDL-C) ratio (MHR) is also presented as a low-cost composite prognostic indicator reflecting the balance between inflammatory and lipid metabolism, as the MHR links the inflammatory cells - monocytes - to the blood lipids - HDL-C. It is known that monocytes have the capacity to migrate into the subendothelial space and take up lipoproteins, and hypercholesterolaemia in particular promotes their faster migration. In contrast, HDL-C molecules have anti-atherosclerotic properties due to their function in the so-called "reverse cholesterol transport", which prevents monocyte activation and recruitment. Thus, MHR is also considered a diagnostic indicator of the presence and severity of CAD [10].

Another cheap reading is the systemic inflammatory response index (SIRI), which is calculated using even three types of white blood cells: neutrophils, lymphocytes and monocytes. Studies have shown that the SIRI has a prognostic value in suspected CAD because it includes neutrophils, which have a tendency to activate macrophages and stimulate further recruitment of monocytes and cytotoxicity, accelerating all stages of atherosclerosis; lymphocytes, which regulate inflammation and therefore have an anti-atherosclerotic effect and monocytes, which promote the inflammatory process that leads to plaque formation, progression and eventual rupture [11]. Li Y. and co-authors conducted a study to assess the association of SIRI with adverse cardiovascular prognosis in patients initially diagnosed with CAD. Their exclusion criteria were quite similar to ours (active tumour or paraneoplastic syndrome, acute infection, severe renal failure, severe hepatic failure, known inflammatory/autoimmune disease, active cerebrovascular disease), but the study included patients with diabetes mellitus [13]. Similarly, Meanwhile, Urbanowicz T et al. attempted to assess the prognostic role of SIRI in the development of CAD in their study. They used few exclusion criteria (acute coronary syndromes, haematological and rheumatic diseases, or history of oncology) and included patients with other comorbidities, such as diabetes mellitus, chronic obstructive pulmonary disease, peripheral arterial disease, renal impairment, and history of stroke [11].

The systemic immune-inflammation index (SII) is presented as another important prognostic marker of CAD. It is calculated by multiplying the platelet count by the neutrophil-to-lymphocyte ratio [12]. The reading reflects the balance between inflammation and immune response. Candemir M. evaluated the association between SII and the severity of coronary atherosclerosis in patients diagnosed with SAP, with the same exclusion criteria as ours, but they also included patients with diabetes mellitus [3]. SII has recently been offered to be used for determination of the severity and prognosis of CAD, as it can provide a more detailed assessment of the inflammatory and immune status of patients with CAD [13, 14].

Our study differs from previous studies in that it included patients with stable angina pectoris (SAP), as we tried to exclude patients with comorbidities that may have affected the laboratory measurements. Our study results should be useful in the future for a more accurate identifying the SAP patients who are at the highest risk of significant coronary artery stenosis and who would benefit from more accurate differential diagnosis. Therefore, this study aimed to investigate the relationship between calculated inflammatory readings based on complete blood test, as an inexpensive and easily measurable indicator, and the severity of coronary atherosclerosis in patients with SAP.

## Materials and methods

### Study population

The retrospective pilot study included 166 eligible patients diagnosed with SAP and examined at the Cardiology Department of Kaunas Clinics between December 2022 and December 2023. The diagnosis of SAP was made according to the criteria set out in the guidelines [15]. Exclusion criteria were aged less than 18 years, history of acute coronary syndrome, previous revascularization (previous coronary artery bypass grafting or percutaneous coronary intervention), peripheral arterial disease, chronic or acute heart failure, reduced left ventricular ejection fraction < 50%, severe valvular heart disease, acute or chronic infections, systemic inflammatory or autoimmune diseases, treatment with glucocorticoids in the last 3 months, recent trauma or major surgery in the last month, as well as oncological, hematological, rheumatic, endocrine diseases and hepatic or kidney failure (glomerular filtration rate < 60 mL/min.).

All these patients underwent invasive coronary angiography (CA) or multi-slice computed tomographic coronary angiography (CCTA), or both, to assess coronary artery disease and its severity. This study was retrospective, so baseline characteristics and results of laboratory and other tests (echocardiography, CA, CCTA) were obtained from the hospital’s electronic medical records: age, sex, body mass index (BMI), heart rate, systolic and diastolic blood pressure (BP), as well as comorbidities such as arterial hypertension (AH), dyslipidemia, and risk factors such as smoking, obesity, early family history of cardiovascular disease, and use of medication (beta-blocker, statin, ezetimibe, angiotensin-converting enzyme (ACE) inhibitor or angiotensin II receptor blocker (ARB), calcium channel blocker (CaCB), mineralocorticoid receptor antagonist (MRA), antiplatelet agent (aspirin), trimetazidine or ranolazine).

Coronary artery disease (CAD) refers to atherosclerotic lesions in the coronary arteries, and SAP is one of the symptoms experienced by people with CAD, which is characterized by a predictable feeling of chest discomfort or pain on exertion, which is alleviated with rest or with the use of medications [15]. In addition, symptoms of atypical angina, characterized by shortness of breath, abdominal distension, gas, abdominal pain, burning or tenderness in the back, shoulders, arms or jaw, more common in women, were assessed and differentiated from other conditions which may mimic symptoms similar to those of stable angina, such as gastroenterological pathology (for example, gastro-esophageal reflux disease), musculoskeletal conditions (for example, straining of the muscles of the chest wall), lung disease, and anxiety or panic attacks. Patients with a history of acute coronary syndrome and previous revascularization, as mentioned before, were excluded.

All the investigations were approved and conducted in accordance with the guidelines of the local Bioethics Committee and adhered to the principles of the Declaration of Helsinki and Title 45, U.S. Code of Federal Regulations, Part 46, Protection of Human Subjects (revised 15 January 2009, effective 14 July 2009). The study was approved by the Regional Bioethics Committee at the Lithuanian University of Health Sciences (No.: BE-2-132, 2021.12.16).

### Laboratory Measurements

Laboratory blood samples were taken before CA or CCTA. The results were obtained from patient histories: complete blood counts, total cholesterol, high-density lipoprotein cholesterol (HDL-C), low-density lipoprotein cholesterol (LDL-C), triglycerides, apolipoprotein B (apoB), lipoprotein (a) (Lp(a)), as well as high-sensitivity C-reactive protein (hs-CRP) and uric acid. The following parameters were analysed from the total blood count: leukocytes (neutrophils, lymphocytes, monocytes, eosinophils), erythrocytes, platelets, and mean platelet volume (MPV), and the ratio of these parameters calculated: PLR was calculated by dividing platelet count by lymphocyte count, NLR – by dividing neutrophil count to lymphocyte count, NMR as the ratio of neutrophil count to monocyte count and LMR - the ratio of lymphocyte count to monocyte count, MLR by dividing monocyte count to lymphocyte count, while MHR – was calculated as the ratio of monocytes to high-density lipoproteins, SIRI was calculated as MLR multiplied by neutrophils and SII was calculated as NLR multiplied by platelets.

Arterial hypertension was defined as systolic and/or diastolic blood pressure ≥ 140 and/or 90 mmHg, respectively. The dyslipidaemia group consisted of patients with total cholesterol concentration greater than 5 mmol/l or LDL-C concentration greater than 3 mmol/l or triglycerides concentrations greater than 1.7 mmol/l or HDL-C less than 1 mmol/l for men, <1.2 mmol/l for women, as well as patients who were already taking a statin, even though their LDL-C and triglycerides concentrations had already been lowered to within the normal limits. Smoking status was defined as current tobacco use. An early family history of cardiovascular disease was defined as early cardiovascular disease in first-degree relatives (age <55 years in men and <65 years in women).

### Instrumental examination

We analysed the following transthoracal echocardiography data from patient history: left ventricular measurements, such as left ventricular end-diastolic diameter and index (LVEDD, LVEDDi), thickness of intraventricular septal (IVS) and left ventricular posterior wall (LVPW), left ventricular mass (LVM) and left ventricular mass index (LVMi), and relative left ventricular wall thickness (RWT), as well as left ventricular ejection fraction (LVEF), left atrial diameter (LA), right atrial and right ventricular diameter (RA, RV), tricuspid annular systolic velocity (RV S’), pulmonary artery systolic pressure (PASP). Also left ventricular filling pressure measurements such as peak early diastolic velocity (E), late diastolic velocity (A), E/A ratio to assess the degree of left ventricular diastolic dysfunction. Mitral annular early diastolic velocity (e′) and late diastolic velocity were measured in the septal (e′_sep_) and lateral (e′_lat_) mitral annulus and the E/e′ ratio was calculated.

For patients with a low or intermediate probability of coronary artery disease, CCTA was selected based on the ESC 2019 guidelines for chronic coronary syndromes [15]. CCTA scans were performed on CT scanner (Canon Aquilion One Genesis) with a minimum of 64 slices. Pre-treatment with nitroglycerin was added according to the recommendations. If the patients’ pre-CCTA heart rate was higher than 60 bpm, heart rate control drugs such as beta-blockers were administered (sinus node I(f) channel inhibitor (ivabradine) was given to patients who could not take beta-blockers due to any contraindication).

Analysis of the scans was performed using a Vitrea workstation. Initially, the images were reconstructed at 75% of the cardiac cycle, with a thickness of 0.5 mm and 0.3 mm, respectively. The spatial resolution was 0.33 mm. In the presence of motion artefacts, additional reconstructions were performed at different time points of the R-R interval. CCTA scans were analyzed on site by a qualified radiologist and patients’ medical histories were concealed.

To determine the severity of coronary stenosis, CAD severity was categorized according to the Coronary Artery Disease Reporting and Data System (CAD-RADS) classification [31]. The CAD-RADS classification consisted of the following variants: CAD-RADS 0 - no plaque or stenosis, CAD-RADS 1 - minimal stenosis: 1 to 24%, CAD-RADS 2 - mild stenosis: 25 to 49%, CAD-RADS 3 - moderate stenosis: 50 to 69%, CAD-RADS 4 - severe stenosis: 70 to 99%, or a left main ≥ 50%, or three-vessel obstructive disease ≥ 70%, CAD-RADS 5 - at least one completely occluded coronary artery.

As recommended in the aforementioned ESC 2019 guidelines on chronic coronary syndromes [15], CA was performed in patients with a high clinical probability and severe symptoms that could not be treated with medication, or if obstructive CAD was suspected on CCTA scan.

CA was performed using standard Judkins methods through femoral artery or radial artery. Coronary angiograms were analyzed by experienced interventional cardiologist who were not informed about the patient’s clinical data. Coronary artery stenosis was judged by visual assessment. To assess the degree of atherosclerotic stenosis, the percentage degree of luminal narrowing was calculated by using the following formula: diameter of the normal vessel in the proximal part of stenotic vessel—diameter of stenotic site)/diameter of the proximal part of stenotic vessel ×100%.

The Gensini scoring system was used to assess the severity of CAD. The Gensini score was calculated for each patient from the coronary arteriogram, assigning a severity score for each form of coronary stenosis based on the degree of luminal narrowing and its geographical significance. The degree of stenosis and the location of the coronary artery lesion were scored as follows: 1 point for ≤ 25% stenosis, 2 points for 26-50% stenosis, 4 points for 51-75% stenosis, 8 points for 76-90% stenosis, 16 points for 91-99% stenosis, and 32 points for complete occlusion. Each lesion score was multiplied by a factor to take into account the importance of the lesion’s position in the coronary circulation: 5 for the proximal segment of the circumflex artery, 1.5 for the middle segment of the left anterior descending coronary artery, 1.0 for the right coronary artery, the distal segment of the left anterior descending coronary artery, the posterolateral artery and the posterolateral artery, and 0.5 for the other segments. Finally, the Gensini score was calculated by summing the scores of the individual coronary segments [16]. All patients diagnosed with SAP underwent tests to assess coronary artery disease.

### Patients’ grouping

After CA or CCTA, or both, patients were divided into two groups: patients without coronary stenosis (patients without CAD group) and patients with any coronary stenosis (patients with CAD group). Most patients were diagnosed with CAD (87.95%, n=146).

Patients were divided into three ways: 1) according to CAD presence or absence; 2) according to Gensini score; 3) according to CAD-RADS score.

Patients who underwent CA were divided into three groups according to a Gensini score designed to assess the relationship between CAD and its severity. The first group consisted of patients with a Gensini score between 0 and 11 (n=44), the second group consisted of patients with a Gensini score between 12 and 35 (n=36), and the third group consisted of patients with a Gensini score above 35 (n=27).

Six groups are used according to the CAD-RADS classification: CAD-RADS 0 - no plaque or stenosis, CAD-RADS 1 - minimal stenosis: 1 to 24%, CAD-RADS 2 - mild stenosis: 25 to 49% and CAD-RADS 3 - moderate stenosis: 50 to 69%, CAD-RADS 4 - severe stenosis: 70 to 99% or left main ≥ 50% or three-vessel obstructive disease ≥ 70%, CAD-RADS 5 - at least one completely occluded coronary artery [31].

Our patients who underwent CCTA were divided into groups according to the CAD-RADS classification, combining them into four groups: the first group consisted of patients with no coronary stenosis (CAD-RADS 0; n=13), the second group consisted of patients with minimal and mild stenosis (CAD-RADS 1 and 2; n=26), the third group consisted of patients with moderate stenosis (CAD-RADS 3; n=21), and finally, the fourth group consisted of patients with significant stenosis (CAD-RADS 4 and 5; n=39).

### Statistical analysis

Statistical analysis was carried out using the program IBM-SPSS Statistics 29.0. Normality tests (Kolmogorov-Smirnov or Shapiro-Wilk) were used to test whether quantitative variables are normally distributed, given the sample size. Continuous variables are reported using means and standard deviation or medians (minimum and maximum values), depending on the distribution model. Categorical variables are presented as numbers and percentages.

Quantitative variables with normal and abnormal distributions were compared using the Student’s t-test and the Mann-Whitney U test. Categorical variables were compared using the chi-square test. The association between SII and the severity of CAD and other variables was assessed using Spearman’s rank correlation coefficient. The Kruskal-Wallis test was used to assess differences in variables between three or more groups. A two-sided p < 0.05 was considered statistically significant for all comparisons.

## Results

### 1. Baseline characteristics of the patients

A total of 166 patients were investigated in this study. The majority of the patients underwent CA (71.69% (n=119)) and more than half of all subjects underwent CCTA (59.64% (n=99)). More than a quarter of patients (27.71% (n=46)) had only one CCTA test, while almost a third of patients (31.93% (n=53)) had both tests to assess coronary atherosclerosis. More than a third of patients underwent CA test (39.76% (n=66)). The majority of patients had coronary atherosclerosis – 87.95% (n=146). Less than half of these patients needed coronary revascularisation: percutaneous coronary intervention was performed in 38% (n=63), while 3% (n=5) required coronary artery bypass grafting.

Patients diagnosed with CAD were statistically significantly older *(p=0.007)* and had higher systolic blood pressure *(p=0.034)* compared to patients without CAD (Table 1). Aspirin use were statistically significantly more common in patients with CAD *(p=0.047*, respectively (Table 2).

**Table 1.** Baseline characteristics of patient’ groups according to CAD presence.

|  | **Patients with CAD**<br>**(n=146)** | **Patients without CAD**<br>**(n=20)** | ***p* value** |
| --- | --- | --- | --- |
| **Age, years** | 60.5 (38-85) | 53 (34-79) | ***0.007*** |
| **Female gender, n (%)** | 60 (41.1) | 10 (50) | *0.477* |
| **BMI, kg/m<sup>2</sup>** | 29.18 (19.57-41.03) | 28.55 (22.04-38.06) | *0.240* |
| **Smoking, n (%)** | 43 (29.7) | 5 (25) | *0.796* |
| **Obesity, n (%)** | 59 (41.5) | 6 (30) | *0.610* |
| **Arterial hypertension, n (%)** | 125 (85.6) | 15 (75) | *0.495* |
| **Early family history of cardiovascular disease, n (%)** | 25 (19.4) | 1 (5) | *0.201* |
| **Systolic BP, mmHg** | 140 (116-185) | 133 (100-180) | ***0.034*** |
| **Diastolic BP, mmHg** | 88 (64-120) | 83.5 (70-110) | *0.582* |
| **Heart rate, bpm** | 68 (50-100) | 70 (56-83) | *0.496* |
| **Uric acid, $\mu\text{mol}$** | 334.5 (154-627) | 345 (180-491) | *0.862* |

*Values are medians, with minimum and maximum values in parentheses or numbers with percentages in parentheses. CAD, coronary artery disease; BMI, body mass index; BP, blood pressure; bpm, beats per minute. Significant p values are in bold. The dyslipidaemia group consisted of patients with total cholesterol concentration greater than 5 mmol/l or LDL-C concentration greater than 3 mmol/l or triglycerides concentrations greater than 1.7 mmol/l or HDL-C less than 1 mmol/l for men, <1.2 mmol/l for women, as well as patients who were already taking a statin, even though their LDL-C and triglycerides concentrations had already been lowered to within the normal limits.*

**Table 2.** Medication use in patient’ groups according to CAD presence.

|  | **Patients with CAD**<br>**(n=146)** | **Patients without CAD**<br>**(n=20)** | **p value** |
| --- | --- | --- | --- |
| **Statin, n (%)** | 78 (53.4) | 7 (35) | *0.154* |
| **Ezetimibe, n (%)** | 7 (4.8) | 0 (0) | *0.317* |
| **ACE inhibitor/ARB, n (%)** | 97 (66.4) | 10 (50) | *0.212* |
| **Beta-blocker, n (%)** | 91 (62.3) | 10 (50) | *0.333* |
| **Calcium channel blocker, n (%)** | 34 (23.3) | 4 (20) | *0.743* |
| **MRA, n (%)** | 9 (6.2) | 0 (0) | *0.602* |
| **Aspirin, n (%)** | 39 (27.3) | 1 (5) | **0.047** |
| **Ranolazine, n (%)** | 7 (4.8) | 1 (5) | *0.895* |
| **Trimetazidine, n (%)** | 11 (7.5) | 1 (5) | *0.805* |

*Values are medians, with minimum and maximum values in parentheses or numbers with percentages in parentheses. CAD, coronary artery disease; ACE, angiotensin-converting enzyme; ARB, angiotensin II receptor blocker; MRA, mineralocorticoid receptor antagonist. Significant p values are in bold.*

A comparison of baseline characteristics and medication use of the patients ranked according to the Gensini score is shown in Tables 3 and 4. No significant differences were found.

**Table 3.**
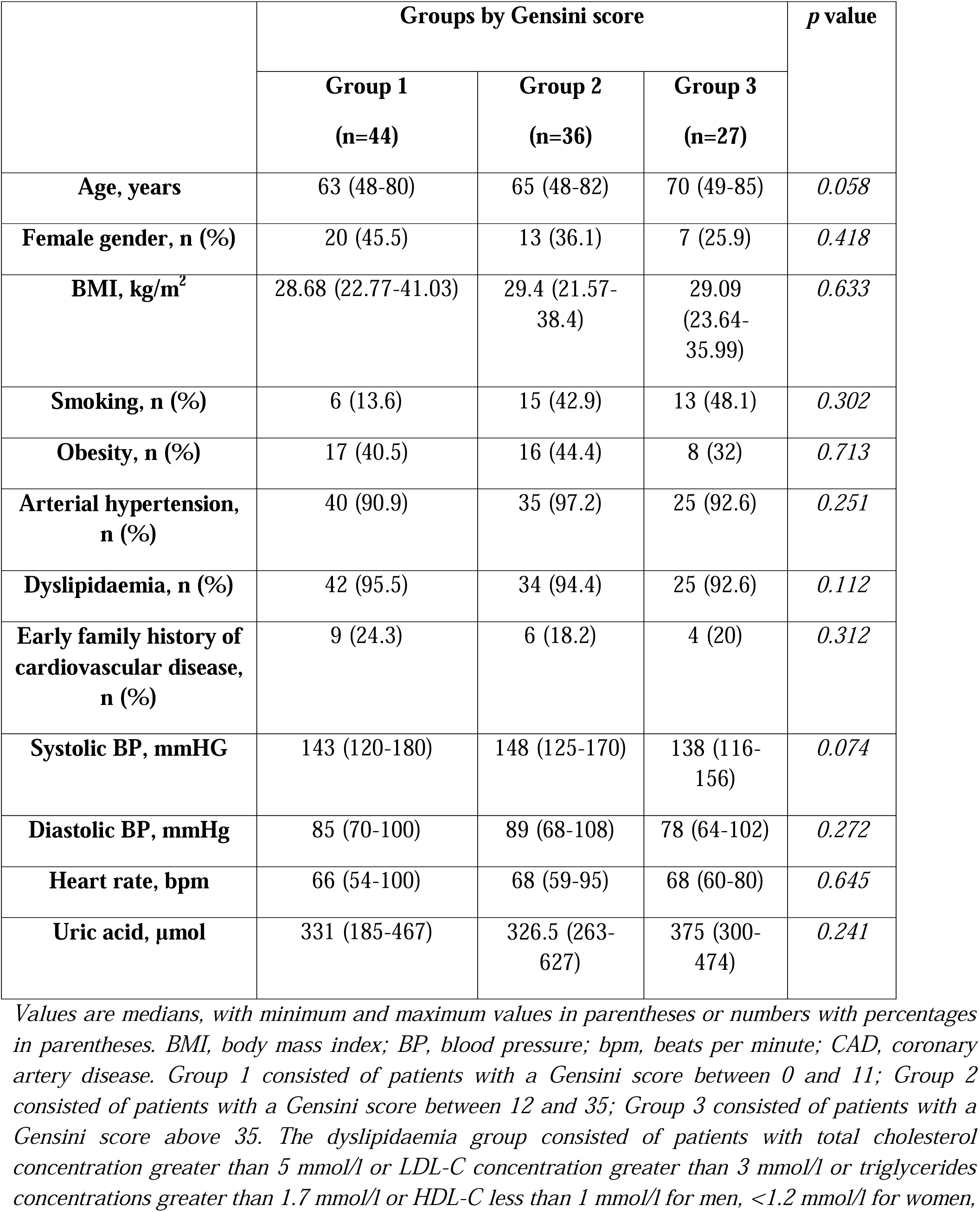
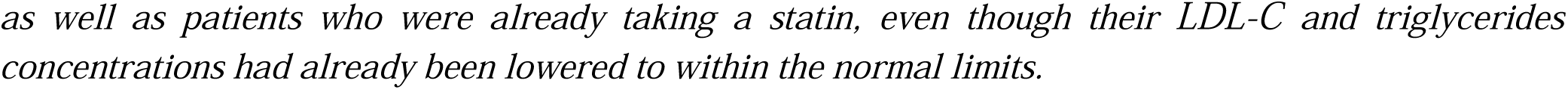
Baseline characteristics of patients with CAD grouped according to Gensini score.

|  | Groups by Gensini score |  |  | *p* value |
| --- | --- | --- | --- | --- |
|  | Group 1<br>(n=44) | Group 2<br>(n=36) | Group 3<br>(n=27) |  |
| **Age, years** | 63 (48-80) | 65 (48-82) | 70 (49-85) | 0.058 |
| **Female gender, n (%)** | 20 (45.5) | 13 (36.1) | 7 (25.9) | 0.418 |
| **BMI, kg/m<sup>2</sup>** | 28.68 (22.77-41.03) | 29.4 (21.57-38.4) | 29.09 (23.64-35.99) | 0.633 |
| **Smoking, n (%)** | 6 (13.6) | 15 (42.9) | 13 (48.1) | 0.302 |
| **Obesity, n (%)** | 17 (40.5) | 16 (44.4) | 8 (32) | 0.713 |
| **Arterial hypertension, n (%)** | 40 (90.9) | 35 (97.2) | 25 (92.6) | 0.251 |
| **Dyslipidaemia, n (%)** | 42 (95.5) | 34 (94.4) | 25 (92.6) | 0.112 |
| **Early family history of cardiovascular disease, n (%)** | 9 (24.3) | 6 (18.2) | 4 (20) | 0.312 |
| **Systolic BP, mmHG** | 143 (120-180) | 148 (125-170) | 138 (116-156) | 0.074 |
| **Diastolic BP, mmHg** | 85 (70-100) | 89 (68-108) | 78 (64-102) | 0.272 |
| **Heart rate, bpm** | 66 (54-100) | 68 (59-95) | 68 (60-80) | 0.645 |
| **Uric acid, µmol** | 331 (185-467) | 326.5 (263-627) | 375 (300-474) | 0.241 |

*Values are medians, with minimum and maximum values in parentheses or numbers with percentages in parentheses. BMI, body mass index; BP, blood pressure; bpm, beats per minute; CAD, coronary artery disease. Group 1 consisted of patients with a Gensini score between 0 and 11; Group 2 consisted of patients with a Gensini score between 12 and 35; Group 3 consisted of patients with a Gensini score above 35. The dyslipidaemia group consisted of patients with total cholesterol concentration greater than 5 mmol/l or LDL-C concentration greater than 3 mmol/l or triglycerides concentrations greater than 1.7 mmol/l or HDL-C less than 1 mmol/l for men, <1.2 mmol/l for women,*
as well as patients who were already taking a statin, even though their LDL-C and triglycerides concentrations had already been lowered to within the normal limits.

**Table 4.** Medication use in patients with CAD grouped according to Gensini score.

|  | Groups by Gensini score |  |  | *p* value |
| --- | --- | --- | --- | --- |
|  | Group 1<br>(n=44) | Group 2<br>(n=36) | Group 3<br>(n=27) |  |
| **Statin, n (%)** | 25 (56.8) | 24 (66.7) | 20 (74.1) | 0.202 |
| **Ezetimibe, n (%)** | 3 (6.8) | 2 (5.6) | 2 (7.4) | 0.816 |
| **ACE inhibitor/ARB, n (%)** | 31 (70.5) | 30 (83.3) | 20 (74.1) | 0.318 |
| **Beta-blocker, n (%)** | 31 (70.5) | 26 (72.2) | 19 (70.4) | 0.520 |
| **Calcium channel blocker, n (%)** | 11 (25) | 9 (25) | 9 (33.3) | 0.864 |
| **MRA, n (%)** | 3 (6.8) | 3 (8.3) | 1 (3.7) | 0.898 |
| **Aspirin, n (%)** | 12 (27.3) | 17 (51.5) | 10 (37) | 0.461 |
| **Ranolazine, n (%)** | 1 (2.3) | 2 (5.6) | 4 (14.8) | 0.558 |
| **Trimetazidine, n (%)** | 3 (6.8) | 4 (11.1) | 4 (14.8) | 0.515 |

*Values are medians, with minimum and maximum values in parentheses or numbers with percentages in parentheses. ACE, angiotensin-converting enzyme; ARB, angiotensin II receptor blocker; MRA, mineralocorticoid receptor antagonist; CAD, coronary artery disease. Group 1 consisted of patients with a Gensini score between 0 and 11; Group 2 consisted of patients with a Gensini score between 12 and 35; Group 3 consisted of patients with a Gensini score above 35.*

Baseline characteristics and drug usage in the groups according to the CAD-RADS classification, are presented in Tables 5 and 6. Patients with moderate (50 – 69%) or severe (more than 70%) coronary stenosis or occlusion of at least one coronary artery, were statistically significantly older in comparison to patients without coronary stenosis (Group 1), *(p=<0.001).* Patients with mild or minimal (1 – 49%) coronary stenosis were statistically significantly more likely to have arterial hypertension in *comparison* with patients without coronary stenosis (Group 1), *(p=0.002; p=0.004; p=0.023,* respectively).

**Table 5.**
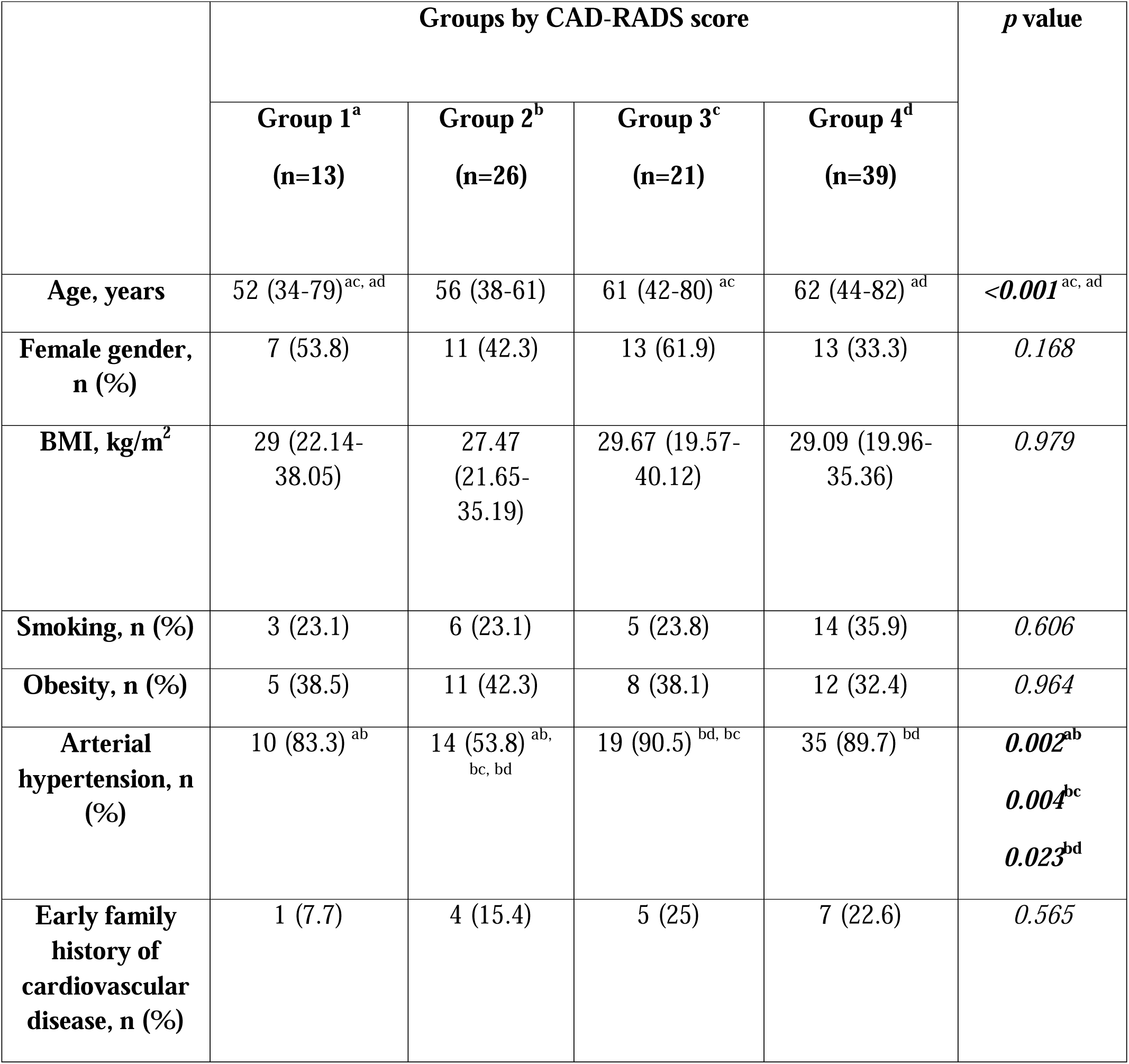
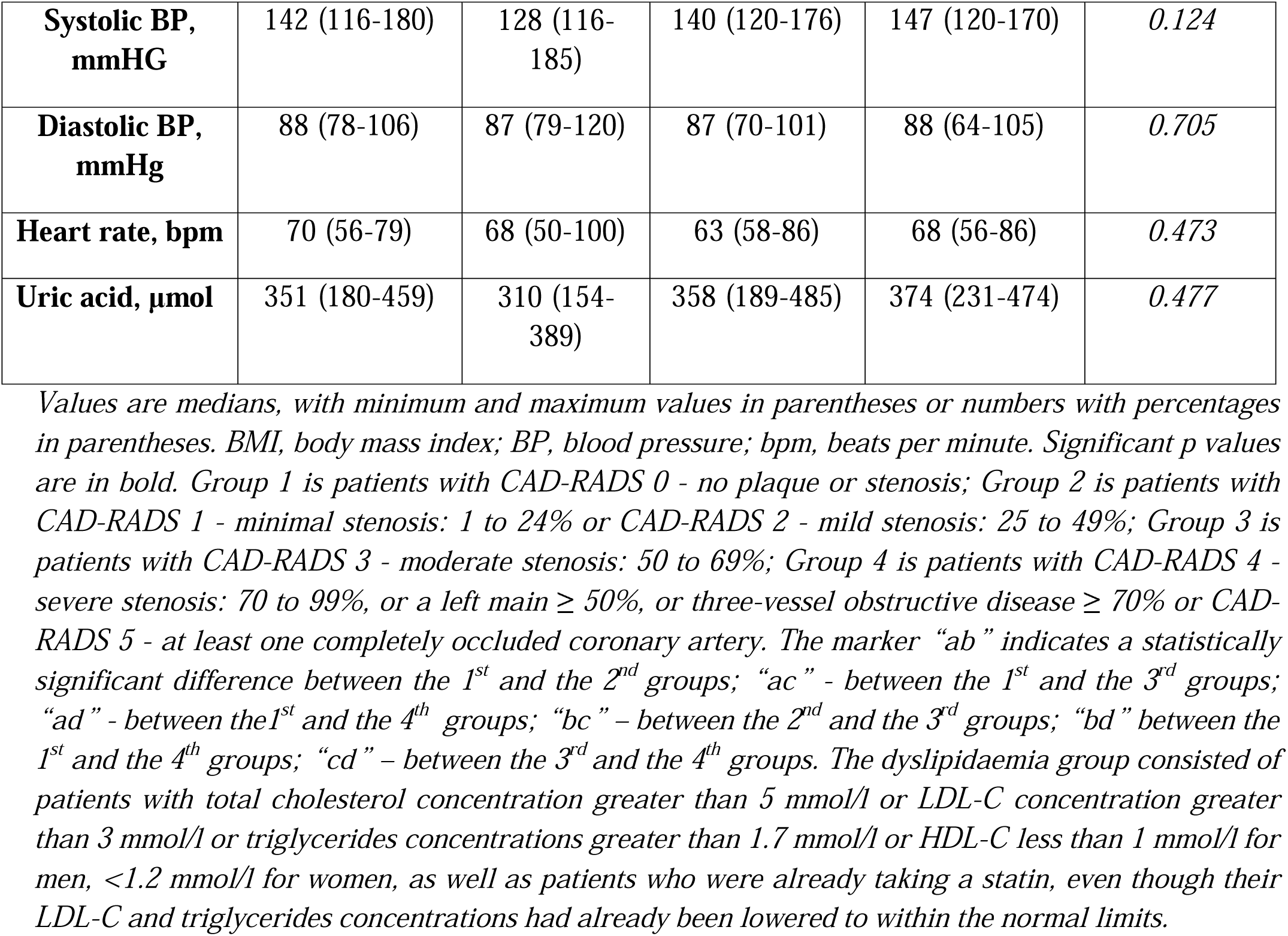
Baseline characteristics of patients grouped according to CAD-RADS score.

Statins were used more frequently in patients with the most severe coronary stenosis (Group 4, Table 6). The differences in usage of ACE inhibitors, beta-blockers and aspirin are presented in Table 6. Usage of other drugs not differed.

**Table 6.**
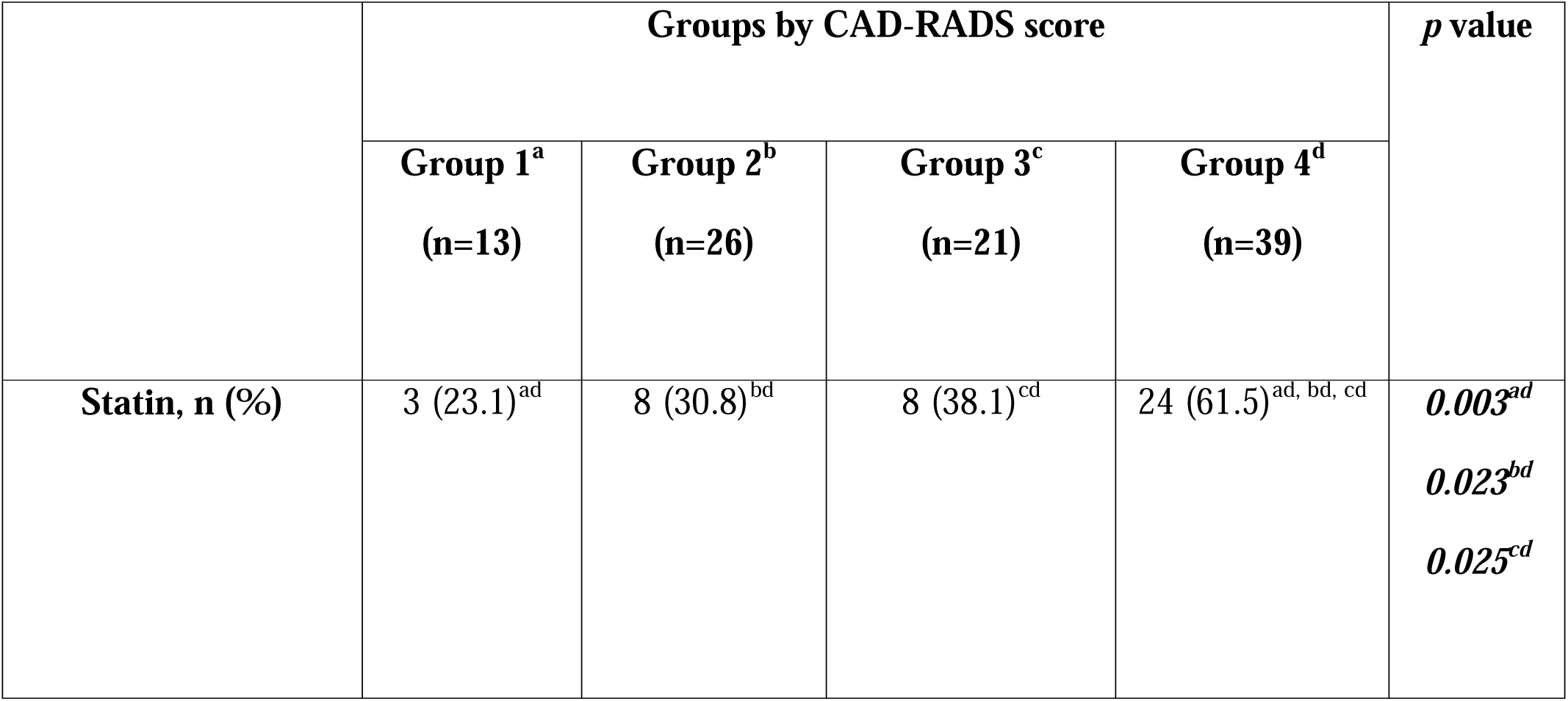
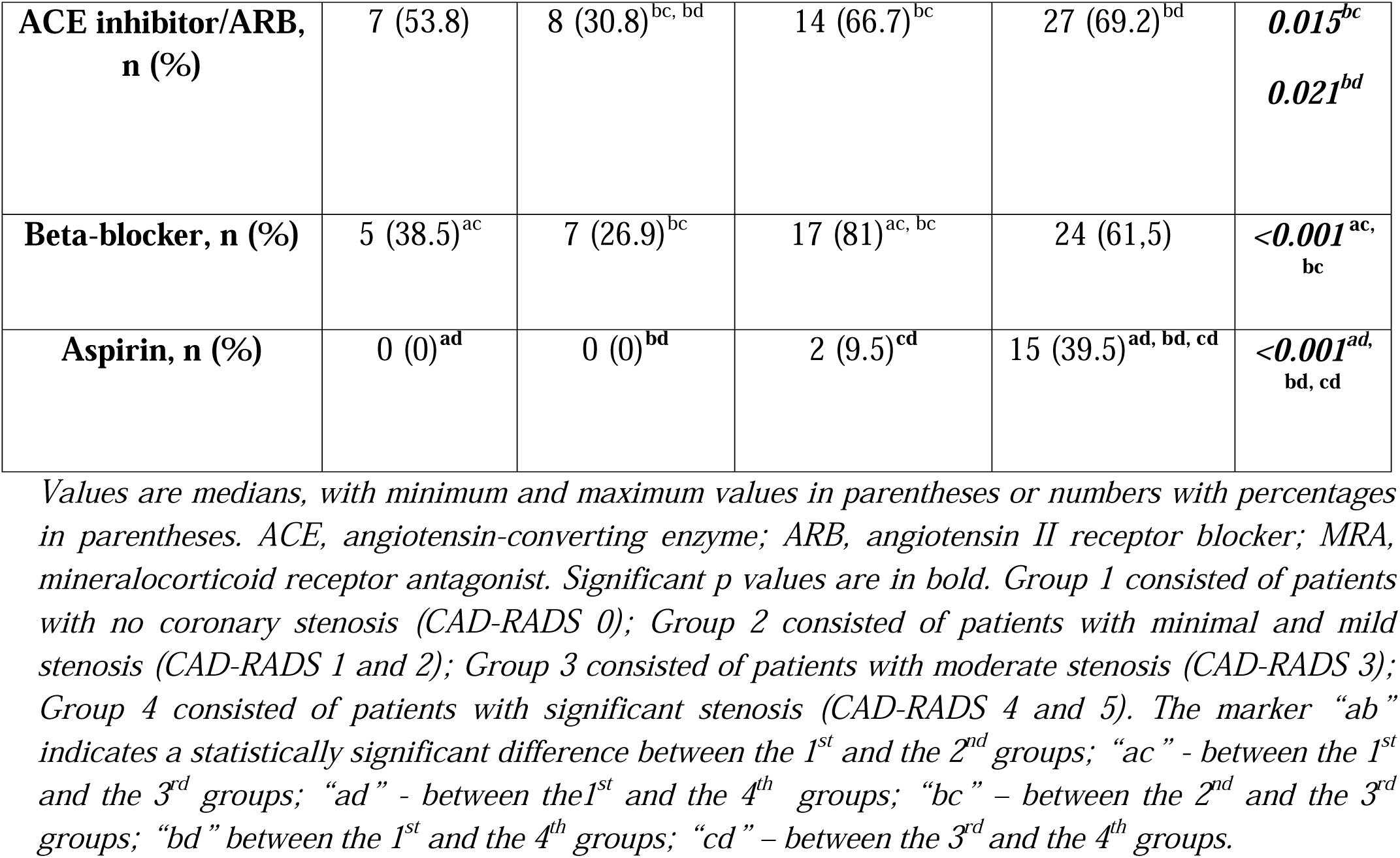
Medication use in patients grouped by CAD-RADS score.

|  | Groups by CAD-RADS score |  |  |  | *p* value |
| --- | --- | --- | --- | --- | --- |
|  | **Group 1<sup>a</sup>**<br>**(n=13)** | **Group 2<sup>b</sup>**<br>**(n=26)** | **Group 3<sup>c</sup>**<br>**(n=21)** | **Group 4<sup>d</sup>**<br>**(n=39)** |  |
| **Statin, n (%)** | 3 (23.1)<sup>ad</sup> | 8 (30.8)<sup>bd</sup> | 8 (38.1)<sup>cd</sup> | 24 (61.5)<sup>ad, bd, cd</sup> | **0.003<sup>ad</sup>**<br>**0.023<sup>bd</sup>**<br>**0.025<sup>cd</sup>** |
| **ACE inhibitor/ARB, n (%)** | 7 (53.8) | 8 (30.8)<sup>bc, bd</sup> | 14 (66.7)<sup>bc</sup> | 27 (69.2)<sup>bd</sup> | **0.015<sup>bc</sup>**<br>**0.021<sup>bd</sup>** |
| **Beta-blocker, n (%)** | 5 (38.5)<sup>ac</sup> | 7 (26.9)<sup>bc</sup> | 17 (81)<sup>ac, bc</sup> | 24 (61.5) | **<0.001<sup>ac, bc</sup>** |
| **Aspirin, n (%)** | 0 (0)<sup>ad</sup> | 0 (0)<sup>bd</sup> | 2 (9.5)<sup>cd</sup> | 15 (39.5)<sup>ad, bd, cd</sup> | **<0.001<sup>ad, bd, cd</sup>** |

Values are medians, with minimum and maximum values in parentheses or numbers with percentages in parentheses. ACE, angiotensin-converting enzyme; ARB, angiotensin II receptor blocker; MRA, mineralocorticoid receptor antagonist. Significant p values are in bold. Group 1 consisted of patients with no coronary stenosis (CAD-RADS 0); Group 2 consisted of patients with minimal and mild stenosis (CAD-RADS 1 and 2); Group 3 consisted of patients with moderate stenosis (CAD-RADS 3); Group 4 consisted of patients with significant stenosis (CAD-RADS 4 and 5). The marker “ab” indicates a statistically significant difference between the 1<sup>st</sup> and the 2<sup>nd</sup> groups; “ac” - between the 1<sup>st</sup> and the 3<sup>rd</sup> groups; “ad” - between the 1<sup>st</sup> and the 4<sup>th</sup> groups; “bc” – between the 2<sup>nd</sup> and the 3<sup>rd</sup> groups; “bd” between the 1<sup>st</sup> and the 4<sup>th</sup> groups; “cd” – between the 3<sup>rd</sup> and the 4<sup>th</sup> groups.

### 2. Lipidogram readings

Total cholesterol, HDL-C, LDL-C, triglycerides, apoB and Lp(a) levels did not differ between patients with and without CAD. Similarly, lipid metabolism readings did not differ between all Gensini score and patients without CAD and CAD-RADS groups.

### 3. Inflammation and total blood count readings

#### 3.1. Inflammation and total blood count readings in patients grouped according to CAD presence

Platelet, erythrocyte count and lymphocyte count and percent, LMR were significantly lower in patients with CAD *(p=0.019; p=0.037; p=0.032; p<0.001; p<0.001,* respectively), while percentage and absolute neutrophils count and NLR were significantly higher in this group compared to patients without CAD *(p=0.002; p=0.021; p<0.001,* respectively). Other markers of systemic inflammation, such as hs-CRP, MLR, SII, SIRI, were statistically significantly higher in the CAD group *(p=0.048; p<0.001; p=0.018; p<0.001,* respectively), (Tables 7 and 8).

**Table 7.** Comparison of inflammation and total blood count readings in patients with and without CAD.

|  | **Patients with CAD**<br>**(n=146)** | **Patients without CAD**<br>**(n=20)** | ***p* value** |
| --- | --- | --- | --- |
| **Hs-CRP, mg/l** | 1.28 (0.12-6.47) | 0.68 (0.12-2.22) | ***0.048*** |
| **Platelet count, x 10<sup>9</sup>/l** | 220 (114-367) | 275 (178-337) | ***0.019*** |
| **Erythrocyte count, x 10<sup>12</sup>/l** | 4.67 (3.40-5.55) | 4.81 (4.11-5.3) | ***0.037*** |
| **Leukocyte count, x 10<sup>9</sup>/l** | 6.4 (3.70-10.27) | 6.27 (4.10-7.60) | *0.148* |
| **Neutrophil count, 10<sup>9</sup>/l** | 3.54 (1.80-7.70) | 3.23 (1.90-4.80) | ***0.021*** |
| **Neutrophil count, %** | 57.50 (32.60 -75.90) | 49.15 (43.30-64.60) | ***0.002*** |
| **Lymphocyte count, 10<sup>9</sup>/l** | 2.00 (1.10-4.12) | 2.28 (1.60-3.30) | ***0.032*** |
| **Lymphocyte count, %** | 31.20 (14.60-48.30) | 37.55 (25.70-46.30) | ***<0.001*** |
| **Monocyte count, 10<sup>9</sup>/l** | 0.50 (0.24-0.88) | 0.50 (0.30-0.68) | *0.052* |
| **Monocyte count, %** | 8.37 (4.20-14.50) | 7.55 (6.20-14.80) | *0.115* |
| **MPV, fl** | 9.00 (6.40-12.80) | 9.15 (6.90-11.90) | *0.407* |

*Values are medians, with minimum and maximum values in parentheses or numbers with percentages in parentheses. CAD, coronary artery disease; hs-CRP, high-sensitivity C-reactive protein; MPV, mean platelet volume. Significant *p* values are in bold.*

**Table 8.** The calculated total blood cell counts readings in patients with and without CAD.

|  | **Patients with CAD**<br>**(n=146)** | **Patients without CAD**<br>**(n=20)** | ***p* value** |
| --- | --- | --- | --- |
| **PLR, 10<sup>9</sup>/l** | 112.61 (48.06-224.29) | 116.09 (81.78-186.25) | *0.585* |
| **NLR, 10<sup>9</sup>/l** | 1.91 (0.67-5.13) | 1.30 (0.96-2.56) | ***<0.001*** |
| **NMR, 10<sup>9</sup>/l** | 7.0 (3.0-15.40) | 6.10 (3.17-10.25) | 0.666 |
| **LMR, 10<sup>9</sup>/l** | 4.00 (1.38-9.50) | 4.37 (2.67-6.61) | **<0.001** |
| **MLR, 10<sup>9</sup>/l** | 0.25 (0.11-0.73) | 0.23 (0.15-0.38) | **<0.001** |
| **SIRI, x 10<sup>9</sup>/l** | 0.93 (0.33-2.57) | 0.70 (0.34-1.14) | **<0.001** |
| **MHR** | 0.38 (0.11-1.0) | 0.33 (0.13-0.85) | 0.062 |

Values are medians, with minimum and maximum values in parentheses or numbers with percentages in parentheses. CAD, coronary artery disease; PLR, platelet to lymphocyte ratio; NLR, neutrophil to lymphocyte ratio; NMR, neutrophil to monocyte ratio; LMR, lymphocyte to monocyte ratio; MLR, monocyte to lymphocyte ratio; SII, systemic immune-inflammation index; SIRI, systemic inflammation response index; MHR, monocyte-to-high-density- lipoprotein ratio. Significant *p* values are in bold.

Patients with CAD had a statistically significantly higher SII compared to patients without the disease (Figure 1).

**Figure 1.**
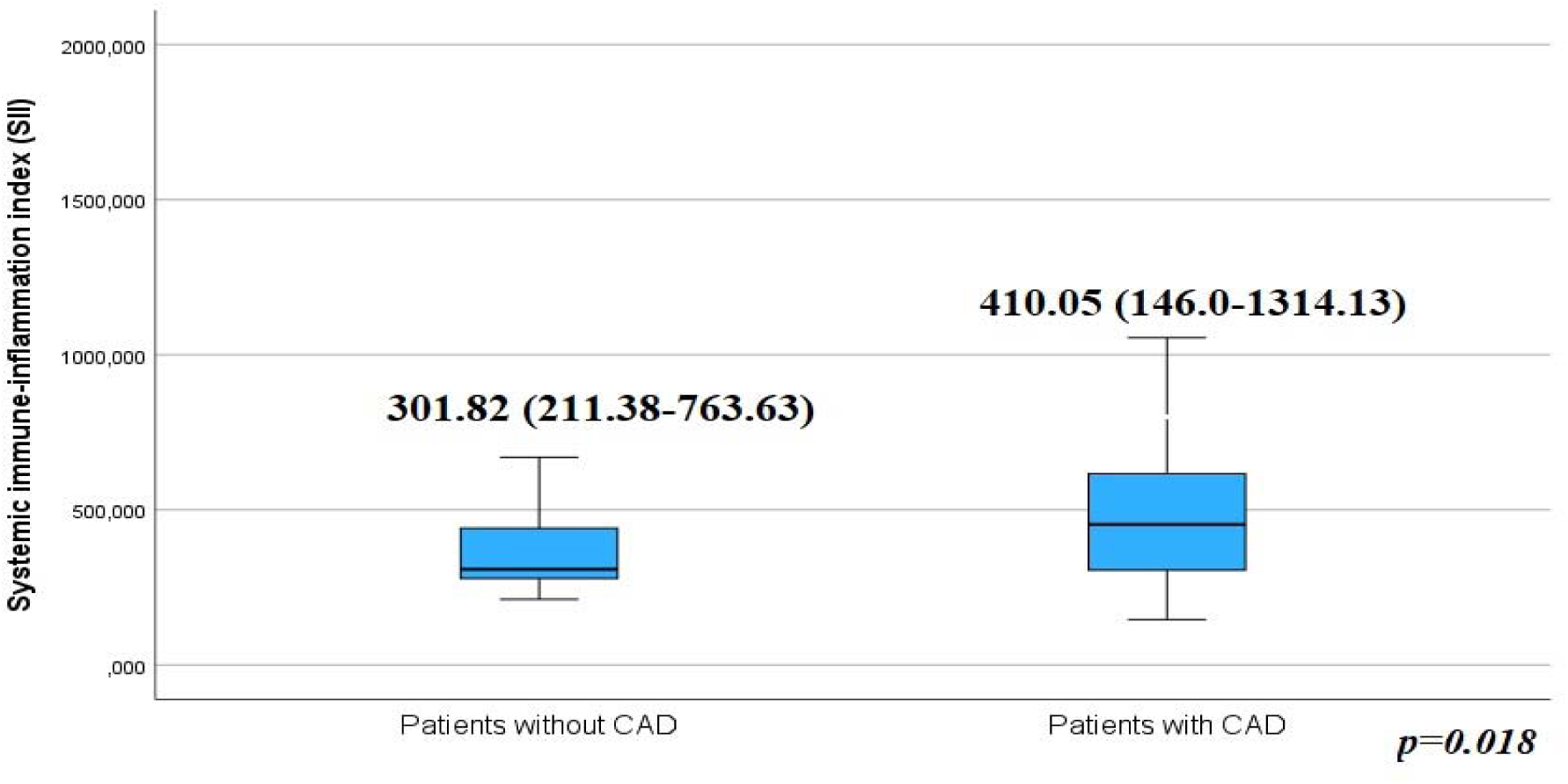
Systemic immune-inflammation index among the groups according to CAD presence. *Values are medians, with minimum and maximum values in parentheses. CAD, coronary artery disease.*

#### 3.2. Inflammation and total blood count readings in patients grouped by Gensini score

Inflammation and total blood levels in patients grouped by Gensini score and in patients without CAD are shown in Tables 9 and 10. Our results showed a tendency to increase in leukocyte count and percentage with the increasing with the, Gensini group (with more severe coronary stenosis). We also found that the lymphocyte count and percent have a tendency to decrease with the increase of the Gensini score group. Monocyte count and percent not differed between the Gensini score groups. The highest percentage of lymphocytes was found in the first Gensini group (with the least significant coronary artery disease) compared to the second and third Gensini groups *(p<0.001,* for all).

**Table 9.**
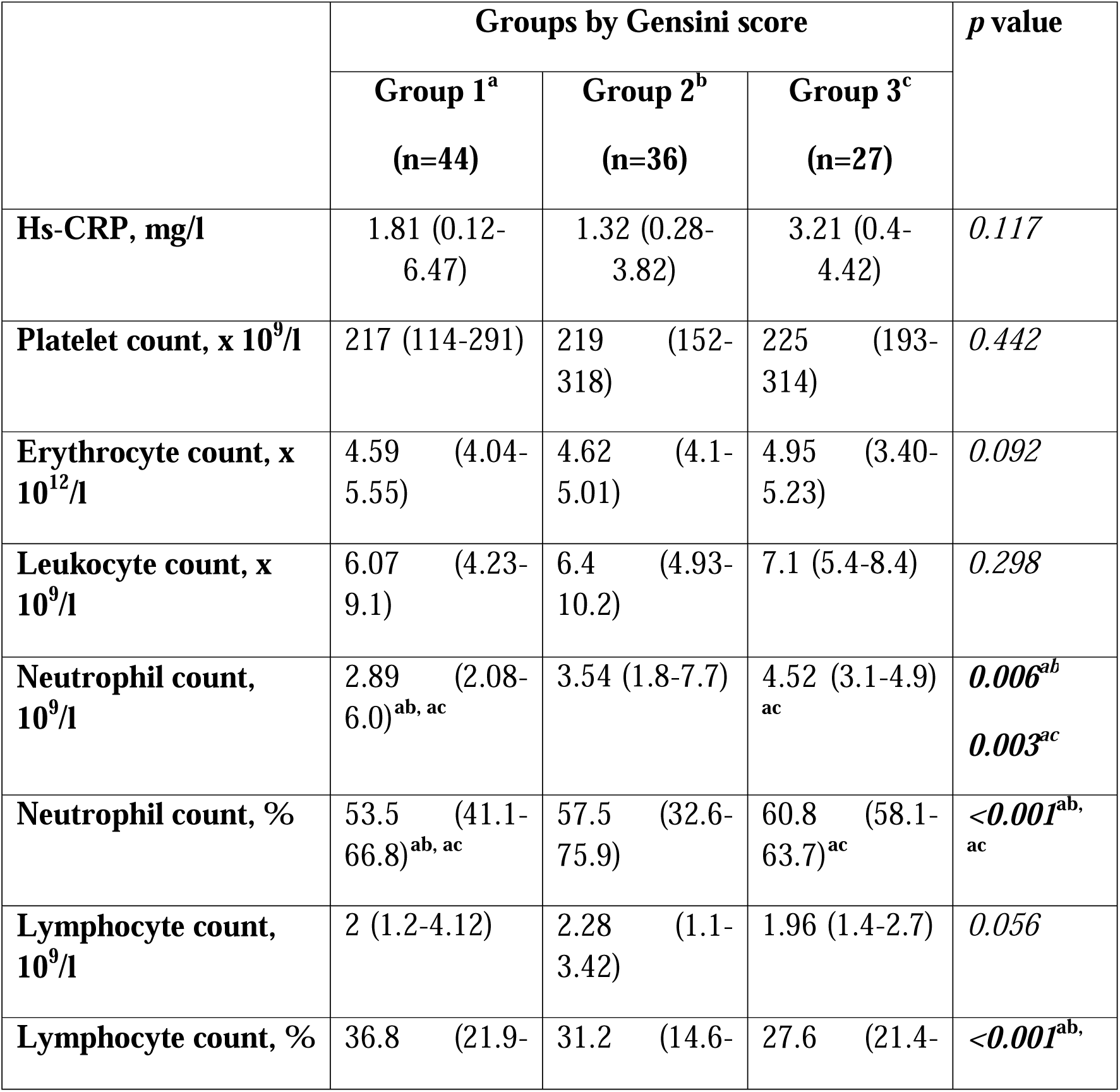
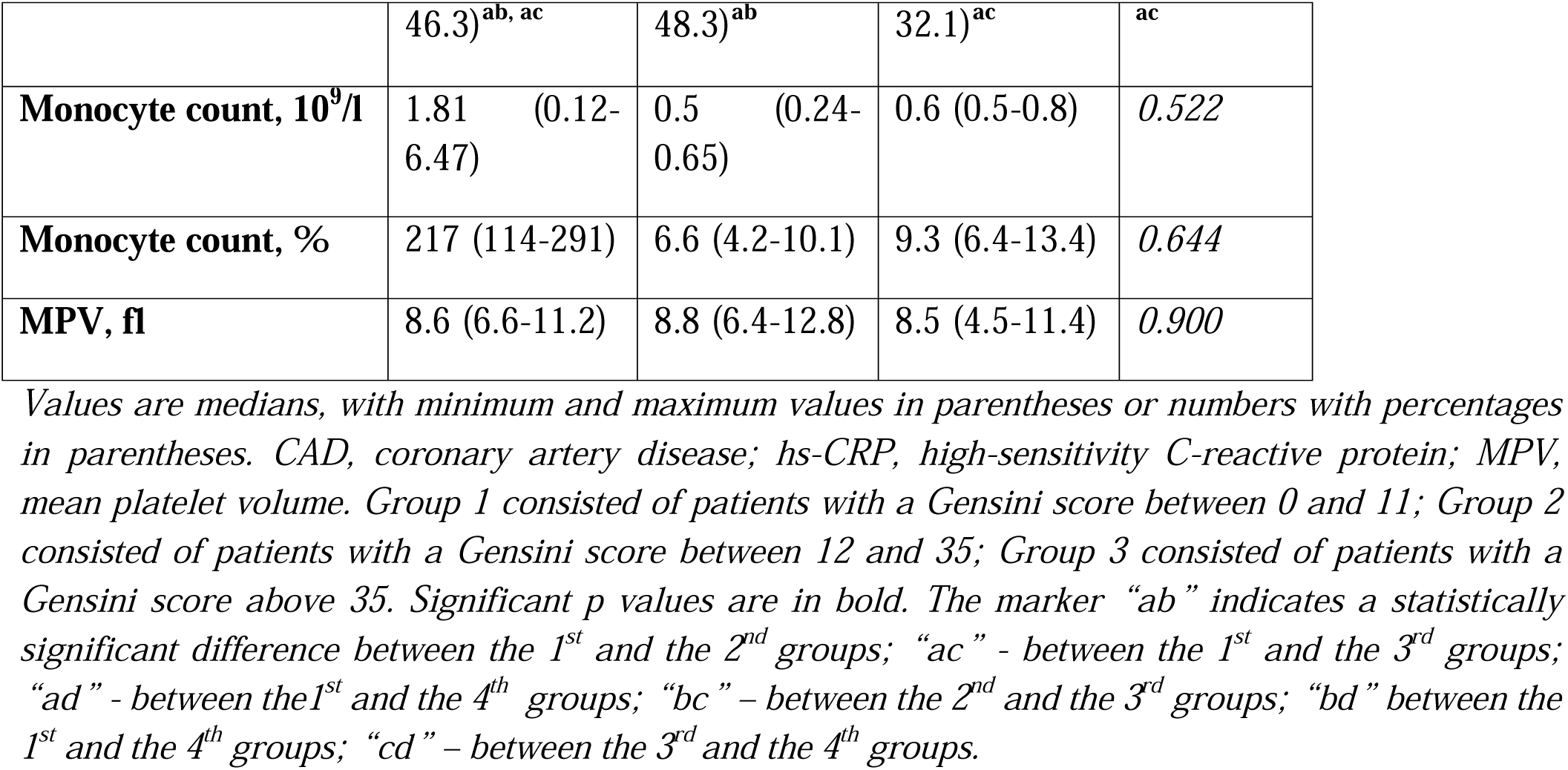
Inflammation and total blood count readings in patients grouped by Gensini score.

**Table 10.** The ratio of total blood cell counts in patients grouped by Gensini score.

|  | Groups by Gensini score |  |  | *p* value |
| --- | --- | --- | --- | --- |
|  | Group 1<sup>a</sup><br>(n=44) | Group 2<sup>b</sup><br>(n=36) | Group 3<sup>c</sup><br>(n=27) |  |
| **PLR, 10<sup>9</sup>/l** | 99.55 (48.06-194)<sup>ac</sup> | 96.07 (63.74-190) | 114.80 (87.73-224.29)<sup>ac</sup> | **0.005<sup>ac</sup>** |
| **NLR, 10<sup>9</sup>/l** | 1.48 (0.89-3.0)<sup>ab, ac</sup> | 1.98 (0.67-5.13)<sup>ab</sup> | 2.21 (1.81-2.93)<sup>ac</sup> | **<0.001<sup>ab, ac</sup>** |
| **NMR, 10<sup>9</sup>/l** | 5.78 (4.05-8.6) | 8.66 (3.0-15.4) | 6.86 (5.16-9.8) | **0.523** |
| **LMR, 10<sup>9</sup>/l** | 4.33 (2.0-5.8)<sup>ac</sup> | 4.6 (2.75-9.5) | 3.14 (1.88-5.4)<sup>ac</sup> | **<0.001<sup>ac</sup>** |
| **MLR, 10<sup>9</sup>/l** | 0.23 (0.17-0.50)<sup>ac</sup> | 0.2 (0.11-0.36)<sup>bc</sup> | 0.32 (0.19-0.53)<sup>ac, bc</sup> | **<0.001<sup>ac, bc</sup>** |
| **SIRI, x 10<sup>9</sup>/l** | 0.72 (0.44-2.4)<sup>ab, ac</sup> | 0.89 (0.33-2.57)<sup>ab</sup> | 1.33 (0.91-2.35)<sup>ac, bc</sup> | **<0.001<sup>ab, ac, bc</sup>** |
| **MHR** | 0.42 (0.22-1) | 0.33 (0.13-0.66) | 0.57 (0.42-0.67) | 0.585 |

Values are medians, with minimum and maximum values in parentheses or numbers with percentages in parentheses. CAD, coronary artery disease; PLR, platelet to lymphocyte ratio; NLR, neutrophil to lymphocyte ratio; NMR, neutrophil to monocyte ratio; LMR, lymphocyte to monocyte ratio; MLR, monocyte to lymphocyte ratio; SII, systemic immune-inflammation index; SIRI, systemic inflammation response index; MHR, monocyte-to-high-density- lipoprotein ratio. Significant *p* values are in bold. Group 1 consisted of patients with a Gensini score between 0 and 11; Group 2 consisted of patients with a Gensini score between 12 and 35; Group 3 consisted of patients with a Gensini score above 35. Significant *p* values are in bold. The marker “ab” indicates a statistically significant difference between the 1<sup>st</sup> and the 2<sup>nd</sup> groups; “ac” - between the 1<sup>st</sup> and the 3<sup>rd</sup> groups; “ad” - between the 1<sup>st</sup> and the 4<sup>th</sup> groups; “bc” – between the 2<sup>nd</sup> and the 3<sup>rd</sup> groups; “bd” between the 1<sup>st</sup> and the 4<sup>th</sup> groups; “cd” – between the 3<sup>rd</sup> and the 4<sup>th</sup> groups.

The NLR, MLR, SII, PLR and SIRI values were the highest in the third Gensini group (with the most severe coronary disease), and the LMR value was the lowest in the third group (with the most severe coronary disease), compared with the first Gensini group (with the least severe coronary disease) and the group without CAD *(p<0.001,* for all).

Hs-CRP has the tendency to increase with the increase of Gensini score group (Table 9). It should be emphasized that the first Gensini group patients had a significantly lower SII, NLR and SIRI than patients in the second and third Gensini groups (with the more severe CAD) (Table 10). (Figure 2).

**Figure 2.**
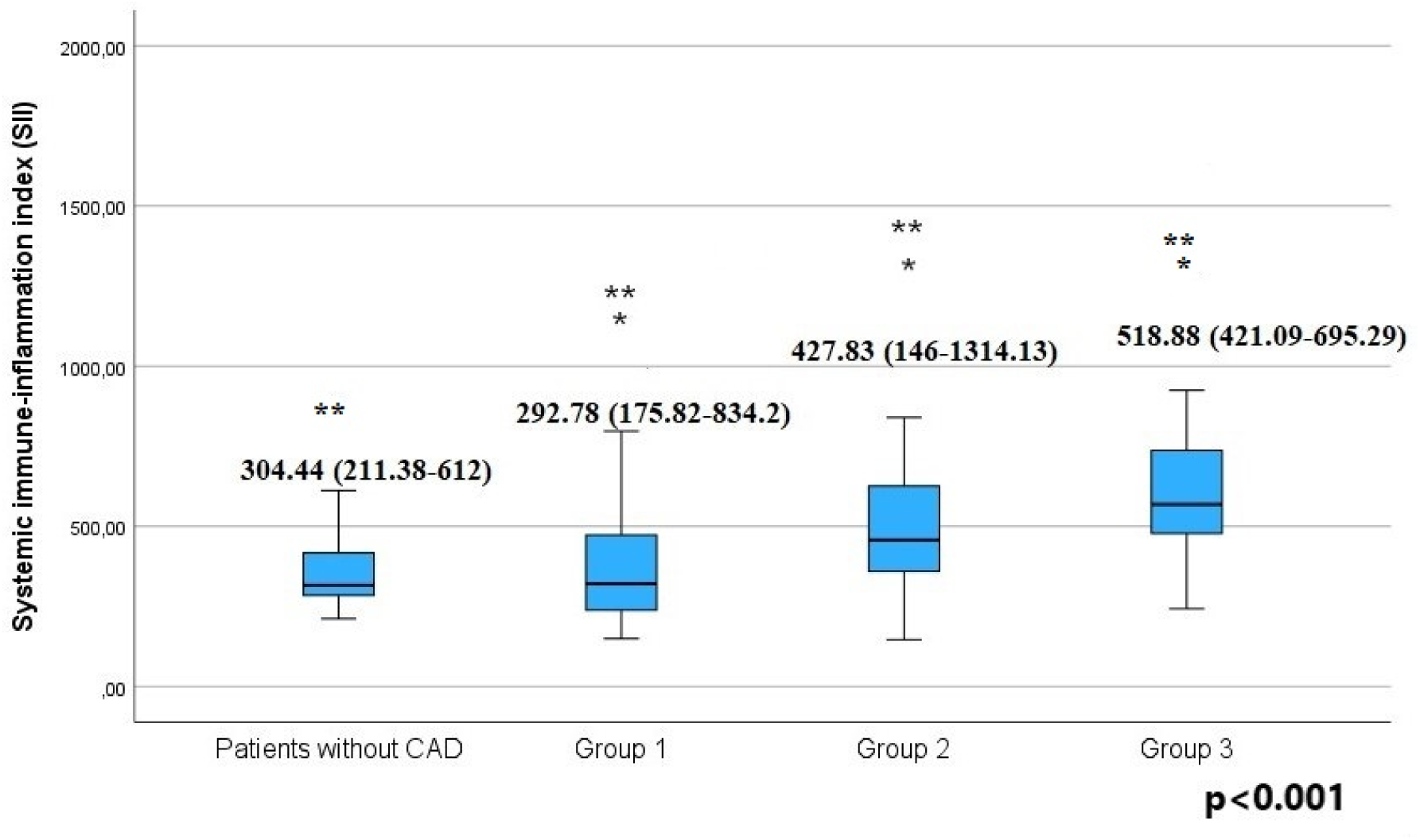
Difference of the systemic immune-inflammation index in the patients grouped according to the Gensini score and patients without CAD. *Values are medians, with minimum and maximum values in parentheses. CAD, coronary artery disease. Group1: patients witha Gensini score between Oand 11; Group 2: patients witha Gensini score between 12 and 35; Group 3: patients with a Gensini score above 35. The marker (*) indicates a statistically significant difference between the first Gensini group with the second and the third Gensini groups. The marker(**) indicates a statistically significant difference between the third Gensini group and the first and the second Gensini groups and patients without CAD*.

#### 3.3 Inflammation and total blood counts readings in patients grouped according to CAD-RADS classification

Platelet, and lymphocyte present had a tendency to decrease with increasing a number of CAD-RADS group (Table 11). Leukocyte count, neutrophil and monocyte count and percent, in opposite, had a tendency to increase. *P*atients without coronary stenosis (Group 1) had lowest NLR, MLR, SIRI, and MHR compared to patients with the most severe coronary stenosis (Group 4) *(p=0.023; p<0.001; p=0.029; p=0.007; p<0.001; p<0.001; p<0.001; p<0.001;* and *p=0.010,* respectively). Meanwhile, lymphocyte percentage and LMR were significantly higher in patients without coronary stenosis (Group 1) compared with patients with the most severe stenosis (Group 4), *(p=0.003; p<0.001)*. LMR was also significantly higher in patients with minimal and mild coronary stenosis (Group 2) compared with patients with the most severe coronary stenosis (Group 4), *(p=0.046).* Our results also showed that patients with the most severe coronary stenosis (Group 4) had significantly higher leucocyte, neutrophil and monocyte counts, as well as SIRI and MHR, compared with patients with minimal and mild coronary stenosis (Group 2), *(p=0.002; p=0.001; p<0.001; p<0.001; p<0.001; p<0.001; p=0.036,* respectively, Table 12).

**Table 11.**
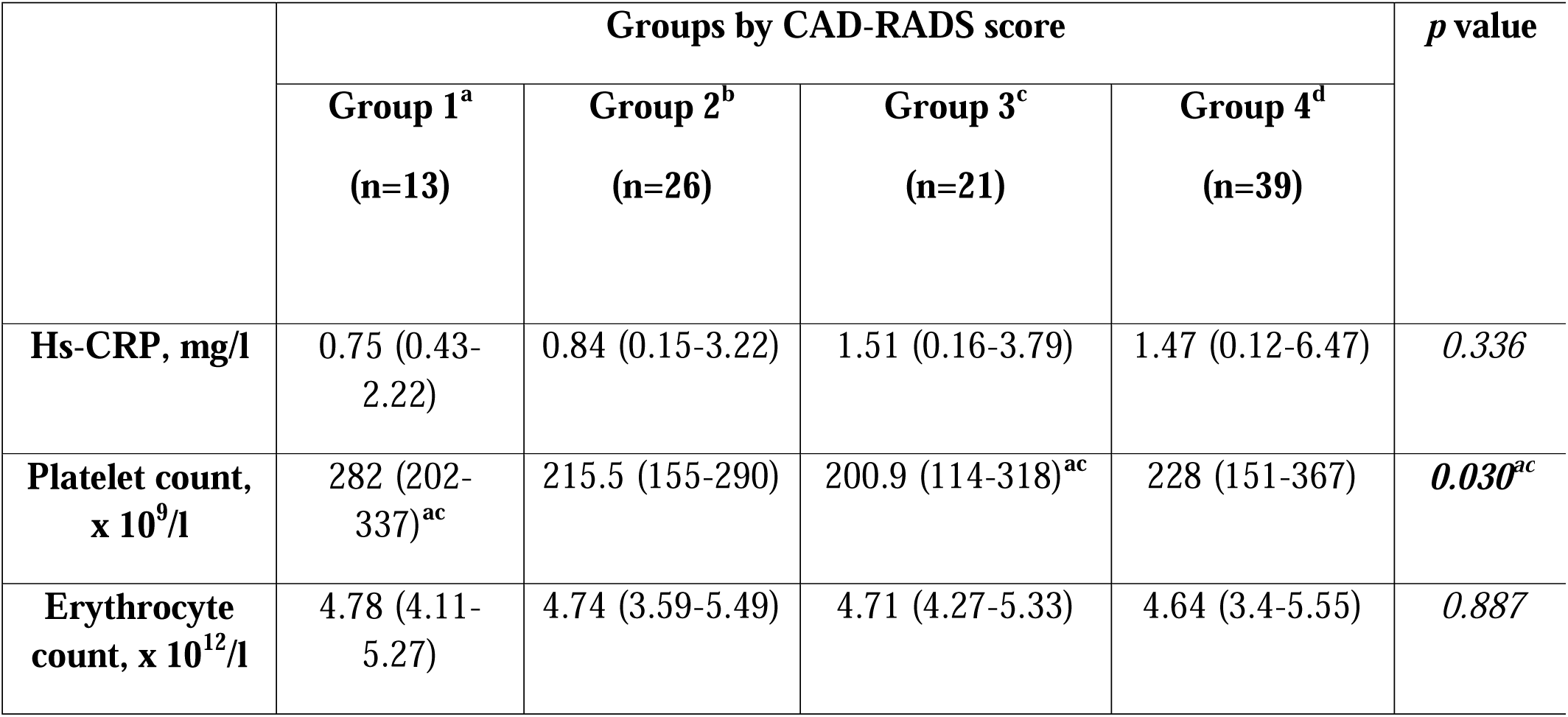
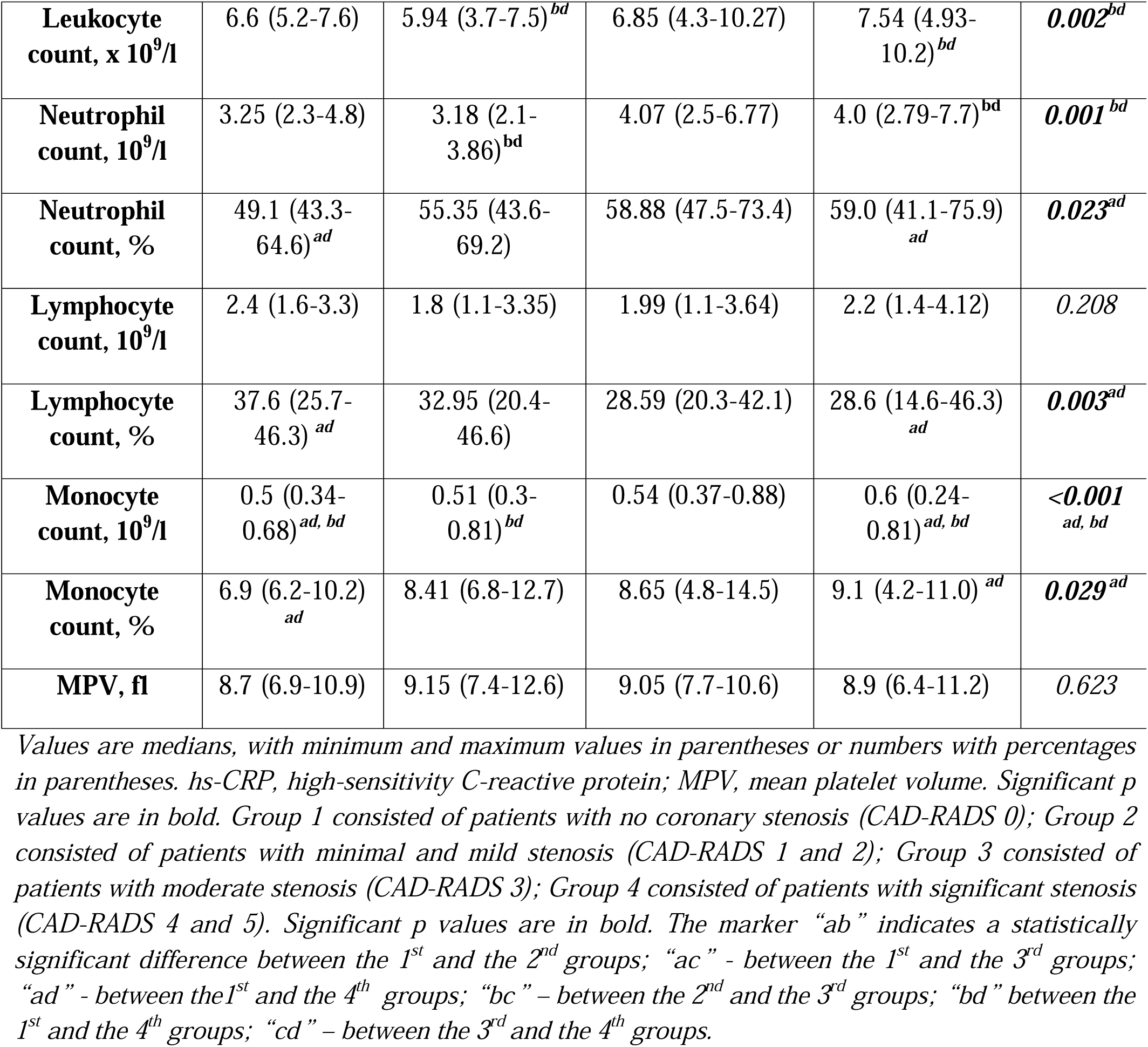
Inflammation and total blood counts readings in patients grouped according to CAD-RADS classification.

|  | Groups by CAD-RADS score |  |  |  | *p* value |
| --- | --- | --- | --- | --- | --- |
|  | Group 1<sup>a</sup><br>(n=13) | Group 2<sup>b</sup><br>(n=26) | Group 3<sup>c</sup><br>(n=21) | Group 4<sup>d</sup><br>(n=39) |  |
| **Hs-CRP, mg/l** | 0.75 (0.43-2.22) | 0.84 (0.15-3.22) | 1.51 (0.16-3.79) | 1.47 (0.12-6.47) | 0.336 |
| **Platelet count, x 10<sup>9</sup>/l** | 282 (202-337)<sup>ac</sup> | 215.5 (155-290) | 200.9 (114-318)<sup>ac</sup> | 228 (151-367) | **0.030<sup>ac</sup>** |
| **Erythrocyte count, x 10<sup>12</sup>/l** | 4.78 (4.11-5.27) | 4.74 (3.59-5.49) | 4.71 (4.27-5.33) | 4.64 (3.4-5.55) | 0.887 |
| **Leukocyte count, x 10<sup>9</sup>/l** | 6.6 (5.2-7.6) | 5.94 (3.7-7.5)<sup>bd</sup> | 6.85 (4.3-10.27) | 7.54 (4.93-10.2)<sup>bd</sup> | **0.002<sup>bd</sup>** |
| **Neutrophil count, 10<sup>9</sup>/l** | 3.25 (2.3-4.8) | 3.18 (2.1-3.86)<sup>bd</sup> | 4.07 (2.5-6.77) | 4.0 (2.79-7.7)<sup>bd</sup> | **0.001<sup>bd</sup>** |
| **Neutrophil count, %** | 49.1 (43.3-64.6)<sup>ad</sup> | 55.35 (43.6-69.2) | 58.88 (47.5-73.4) | 59.0 (41.1-75.9)<sup>ad</sup> | **0.023<sup>ad</sup>** |
| **Lymphocyte count, 10<sup>9</sup>/l** | 2.4 (1.6-3.3) | 1.8 (1.1-3.35) | 1.99 (1.1-3.64) | 2.2 (1.4-4.12) | 0.208 |
| **Lymphocyte count, %** | 37.6 (25.7-46.3)<sup>ad</sup> | 32.95 (20.4-46.6) | 28.59 (20.3-42.1) | 28.6 (14.6-46.3)<sup>ad</sup> | **0.003<sup>ad</sup>** |
| **Monocyte count, 10<sup>9</sup>/l** | 0.5 (0.34-0.68)<sup>ad, bd</sup> | 0.51 (0.3-0.81)<sup>bd</sup> | 0.54 (0.37-0.88) | 0.6 (0.24-0.81)<sup>ad, bd</sup> | **<0.001<sup>ad, bd</sup>** |
| **Monocyte count, %** | 6.9 (6.2-10.2)<sup>ad</sup> | 8.41 (6.8-12.7) | 8.65 (4.8-14.5) | 9.1 (4.2-11.0)<sup>ad</sup> | **0.029<sup>ad</sup>** |
| **MPV, fl** | 8.7 (6.9-10.9) | 9.15 (7.4-12.6) | 9.05 (7.7-10.6) | 8.9 (6.4-11.2) | 0.623 |

Values are medians, with minimum and maximum values in parentheses or numbers with percentages in parentheses. hs-CRP, high-sensitivity C-reactive protein; MPV, mean platelet volume. Significant *p* values are in bold. Group 1 consisted of patients with no coronary stenosis (CAD-RADS 0); Group 2 consisted of patients with minimal and mild stenosis (CAD-RADS 1 and 2); Group 3 consisted of patients with moderate stenosis (CAD-RADS 3); Group 4 consisted of patients with significant stenosis (CAD-RADS 4 and 5). Significant *p* values are in bold. The marker “ab” indicates a statistically significant difference between the 1<sup>st</sup> and the 2<sup>nd</sup> groups; “ac” - between the 1<sup>st</sup> and the 3<sup>rd</sup> groups; “ad” - between the 1<sup>st</sup> and the 4<sup>th</sup> groups; “bc” – between the 2<sup>nd</sup> and the 3<sup>rd</sup> groups; “bd” between the 1<sup>st</sup> and the 4<sup>th</sup> groups; “cd” – between the 3<sup>rd</sup> and the 4<sup>th</sup> groups.

**Table 12.** Comparison of the ratio of total blood cell counts in patients grouped according to CAD-RADS classification.

|  | Groups by CAD-RADS score |  |  |  | *p* value |
| --- | --- | --- | --- | --- | --- |
|  | Group 1<sup>a</sup><br>(n=13) | Group 2<sup>b</sup><br>(n=26) | Group 3<sup>c</sup><br>(n=21) | Group 4<sup>d</sup><br>(n=39) |  |
| **PLR, 10<sup>9</sup>/l** | 120.36<br>(81.78-186.25) | 119.54 (50.15-193.34) | 109.64 (73.08-180.0) | 119.47 (48.06-224.29) | 0.360 |
| **NLR, 10<sup>9</sup>/l** | 1.29 (0.96-2.56)<sup>ad</sup> | 1.66 (0.93-3.28) | 2.07 (1.13-3.59) | 2.09 (0.89-5.14)<sup>ad</sup> | **0.007<sup>ad</sup>** |
| **NMR, 10<sup>9</sup>/l** | 6.45 (4.73-10.25) | 6.57 (3.67-9.0) | 7.05 (4.17-15.27) | 6.86 (4.51-15.4) | 0.937 |
| **LMR, 10<sup>9</sup>/l** | 5.08 (3.68-6.61)<sup>ad</sup> | 3.95 (1.84-5.58)<sup>bd</sup> | 4.17 (1.38-5.71) | 3.17 (2.34-9.5)<sup>ad, bd</sup> | **<0.001<sup>ad</sup>**<br>**0.046<sup>bd</sup>** |
| **MLR, 10<sup>9</sup>/l** | 0.20 (0.15-0.27)<sup>ad</sup> | 0.25 (0.18-0.55) | 0.24 (0.18-0.73) | 0.32 (0.11-0.43)<sup>ad</sup> | **<0.001<sup>ad</sup>** |
| **SII, x 10<sup>9</sup>/l** | 312.29<br>(265.46-763.62) | 342.73 (156.97-674.55) | 432.72 (222.75-876.63) | 453.0 (175.41-1314.13) | 0.113 |
| **SIRI, x 10<sup>9</sup>/l** | 0.65 (0.35-1.14)<sup>ad</sup> | 0.78 (0.53-1.91)<sup>bd</sup> | 1.2 (0.56-2.55) | 1.2 (0.33-2.57)<sup>ad, bd</sup> | **<0.001<sup>ad, bd</sup>** |
| **MHR** | 0.28 (0.13-0.85)<sup>ad</sup> | 0.36 (0.11-0.72)<sup>bd</sup> | 0.4 (0.22-0.65) | 0.48 (0.13-1.0)<sup>ad, bd</sup> | **0.010<sup>ad</sup>**<br>**0.036<sup>bd</sup>** |

Values are medians, with minimum and maximum values in parentheses or numbers with percentages in parentheses. PLR, platelet to lymphocyte ratio; NLR, neutrophil to lymphocyte ratio; NMR, neutrophil to monocyte ratio; LMR, lymphocyte to monocyte ratio; MLR, monocyte to lymphocyte ratio; SII, systemic immune-inflammation index; SIRI, systemic inflammation response index; MHR, monocyte-to-high-density-lipoprotein ratio. Significant *p* values are in bold. Group 1 consisted of patients with no coronary stenosis (CAD-RADS 0); Group 2 consisted of patients with minimal and mild stenosis (CAD-RADS 1 and 2); Group 3 consisted of patients with moderate stenosis (CAD-RADS 3); Group 4 consisted of patients with significant stenosis (CAD-RADS 4 and 5). Significant *p* values are in bold. The marker “ab” indicates a statistically significant difference between the 1<sup>st</sup> and the 2<sup>nd</sup> groups; “ac” - between the 1<sup>st</sup> and the 3<sup>rd</sup> groups; “ad” - between the 1<sup>st</sup> and the 4<sup>th</sup> groups; “bc” – between the 2<sup>nd</sup> and the 3<sup>rd</sup> groups; “bd” between the 1<sup>st</sup> and the 4<sup>th</sup> groups; “cd” – between the 3<sup>rd</sup> and the 4<sup>th</sup> groups.

### 4. Correlations between laboratory, echocardiography, and baseline characteristic readings

#### 4.1. Correlations between echocardiography, total blood count and inflammation readings

Correlations between echocardiography and total blood count readings presented in Table 13. Accordingly, IVS and LVPW thickness weakly correlated with hs-CRP levels *(r=0.360, p<0.001; r=0.256; p=0.021,* respectively). There was a weak correlation between LVM and hs-CRP levels, SIRI and NLR *(r=0.291, p=0.008; r=0.206, p=0.01; r=0.229, p=0.004*, respectively).

**Table 13.** Correlations between echocardiography and total blood count readings.

| Readings | PLT | Neutrophil percentage | Neutrophil count | Lymphocyte percentage | Lymphocyte count |
| --- | --- | --- | --- | --- | --- |
| **LVEF** | $r=0.130$,<br>$p=0.100$ | $r=-0.236$,<br>$p=0.002$ | $r=-0.232$,<br>$p=0.003$ | $r=0.236$<br>$p=0.003$ | $r=0.121$,<br>$p=0.126$ |
| **LVEDD** | $r=-0.248$,<br>$p=0.002$ | $r=0.187$,<br>$p=0.021$ | $r=0.173$<br>$p=0.032$ | $r=-0.239$,<br>$p=0.003$ | $r=-0.160$,<br>$p=0.048$ |
| **IVS thickness** | $r=-0.129$,<br>$p=0.110$ | $r=0.163$,<br>$p=0.043$ | $r=0.139$,<br>$p=0.087$ | $r=-0.112$,<br>$p=0.168$ | $r=-0.036$,<br>$p=0.654$ |
| **LVM** | $r=-0.272$,<br>$p<0.001$ | $r=0.231$,<br>$p=0.004$ | $r=0.179$,<br>$p=0.027$ | $r=-0.218$,<br>$p=0.007$ | $r=-0.123$,<br>$p=0.131$ |
| **LVMi** | $r=-0.261$,<br>$p=0.001$ | $r=0.218$,<br>$p=0.007$ | $r=0.128$,<br>$p=0.115$ | $r=-0.181$,<br>$p=0.025$ | $r=-0.127$,<br>$p=0.119$ |
| **LA diameter** | $r=-0.270$,<br>$p<0.001$ | $r=0.138$,<br>$p=0.094$ | $r=0.126$,<br>$p=0.125$ | $r=-0.171$,<br>$p=0.037$ | $r=-0.098$,<br>$p=0.073$ |
| **LA volume** | $r=-0.350$,<br>$p<0.001$ | $r=0.192$,<br>$p=0.080$ | $r=0.108$,<br>$p=0.329$ | $r=-0.195$,<br>$p=0.075$ | $r=-0.197$,<br>$p=0.073$ |
| **LA volume index** | ***$r=-0.282$,***<br>***$p=0.015$*** | ***$r=0.230$,***<br>***$p=0.049$*** | $r=0.140$,<br>$p=0.234$ | ***$r=-0.242$,***<br>***$p=0.038$*** | $r=-0.211$,<br>$p=0.071$ |
| **$e'_{lat}$** | ***$r=0.221$,***<br>***$p=0.029$*** | ***$r=-0.216$,***<br>***$p=0.033$*** | $r=-0.126$,<br>$p=0.215$ | ***$r=0.232$,***<br>***$p=0.021$*** | $r=0.197$,<br>$p=0.052$ |
| **$e'_{sep}$** | ***$r=0.362$,***<br>***$p<0.001$*** | ***$r=-0.250$,***<br>***$p=0.014$*** | $r=-0.139$,<br>$p=0.177$ | ***$r=0.316$,***<br>***$p=0.002$*** | ***$r=0.273$,***<br>***$p=0.007$*** |
| **$e'$** | ***$r=0.336$,***<br>***$p<0.001$*** | ***$r=-0.249$,***<br>***$p=0.013$*** | $r=-0.153$,<br>$p=0.132$ | ***$r=0.293$,***<br>***$p=0.003$*** | ***$r=0.242$,***<br>***$p=0.016$*** |
| **$E/e'$** | ***$r=-0.228$,***<br>***$p=0.024$*** | $r=0.072$,<br>$p=0.484$ | $r=0.108$,<br>$p=0.292$ | $r=-0.131$,<br>$p=0.198$ | $r=-0.110$,<br>$p=0.281$ |

*PLT, platelets; MPV, mean platelet volume; LVEF, left ventricular ejection fraction; LVEDD, left ventricular end-diastolic diameter; IVS, intraventricular septal; LVM, left ventricular mass; LVMi, left ventricular mass index; LA, left atrium; $e'_{lat}$ , late diastolic velocity were measured in the lateral mitral annulus; $e'_{sep}$ , late diastolic velocity were measured in the septal mitral annulus; $e'$ , average $e'_{sep}$ and $e'_{lat}$ ; $E/e'$ , ratio of peak early diastolic velocity (E) and $e'$ . Significant Spearman's correlations and p-values are in bold.*

RWT weakly correlated with MLR *(r=-0.228, p=0.005)* and LMR *(r=0.228, p=0.005).* LA volume weakly correlated with MLR *(r=0.217, p=0.047)* and LMR *(r=-0.217, p=0.047).* LA volume index weakly correlated with NLR *(r=0.242, p=0.038)* and SIRI *(r=0.244, p=0.036).* NLR weakly correlated with e′_lat_ *(r=-0.243, p=0.016).* No significant correlations were found between RV, RA, tricuspid annular systolic velocity, PASP and total blood count and inflammation readings. Other correlations between echocardiography and calculated readings are presented in Table 14.

**Table 14.** Correlations between echocardiography and total blood count calculated readings.

| **Readings** | **LVEF** | **LVEDD** | **LA diameter** | **$e'$** | **$e'_{sep}$** |
| --- | --- | --- | --- | --- | --- |
| **NLR** | ***$r=-0.230$,***<br>***$p=0.003$*** | ***$r=0.229$,***<br>***$p=0.004$*** | $r=0.149$,<br>$p=0.071$ | ***$r=-0.291$,***<br>***$p=0.004$*** | ***$r=-0.305$,***<br>***$p=0.003$*** |
| **MLR** | ***$r=-0.186$,***<br>***$p=0.018$*** | $r=0.016$,<br>$p=0.842$ | ***$r=0.269$,***<br>***$p<0.001$*** | ***$r=-0.202$,***<br>***$p=0.046$*** | ***$r=-0.302$,***<br>***$p=0.003$*** |
| **LMR** | ***$r=0.186$,***<br>***$p=0.018$*** | ***$r=-0.263$,***<br>***$p=0.001$*** | ***$r=-0.270$,***<br>***$p<0.001$*** | ***$r=0.203$,***<br>***$p=0.045$*** | ***$r=0.302$,***<br>***$p=0.003$*** |
| **SIRI** | r=-0.241,<br>p=0.002 | **r=0.260,**<br>**p=0.001** | **r=0.252,**<br>**p=0.002** | **r=-0.235,**<br>**p=0.020** | **r=-0.304,**<br>**p=0.003** |
| **MHR** | r=-0.113,<br>p=0.179 | **r=0.180,**<br>**p=0.039** | **r=0.217,**<br>**p=0.014** | r=-0.064,<br>p=0.550 | r=-0.125,<br>p=0.248 |

LVEF, left ventricular ejection fraction; LVEDD, left ventricular end-diastolic diameter; LA, left atrium; $e'_{sep}$ , late diastolic velocity were measured in the septal mitral annulus; $e'$ , average $e'_{sep}$ and $e'_{lat}$ ; NLR, neutrophil to lymphocyte ratio; MLR, monocyte to lymphocyte ratio, LMR, lymphocyte to monocyte ratio; NMR, neutrophil to monocyte ratio; PLR, platelet to lymphocyte ratio; SII, systemic immune-inflammation index; SIRI, systemic inflammation response index; MHR, monocyte-to-high-density- lipoprotein ratio; hs-CRP, high-sensitivity C-reactive protein. Significant Spearman's correlations and p-values are in bold.

#### 4.2. Correlations between lipid, and total blood count and inflammation readings

Correlations between total blood count, inflammation and lipidogram showed some low statistically significant correlations (Tables 15 and 16). Hs-CRP levels correlated with neutrophil percentage *(r=0.296, p=0.007)*, neutrophil count *(r=0.298, p=0.007)*, percentage of lymphocytes *(r=-0.279, p=0.011),* value of NLR *(r=0.292, p=0.008)*, NMR *(r=0.260, p=0.018)* and SIRI *(r=0.221, p=0.046)*.

**Table 15.** Correlations between lipid and total blood count readings.

| Readings | Total cholesterol concentration | LDL | HDL | Lp(a) |
| --- | --- | --- | --- | --- |
| **Leukocyte count** | r=-0.079,<br>p=0.347 | r=-0.100,<br>p=0.226 | **r=-0.206,**<br>**p=0.013** | **r=-0.290,**<br>**p=0.012** |
| **Neutrophil count** | r=-0.078,<br>p=0.352 | r=-0.070,<br>p=0.401 | **r=-0.165,**<br>**p=0.049** | r=-0.153,<br>p=0.194 |
| **Monocyte count** | **r=-0.207,**<br>**p=0.013** | **r=-0.162,**<br>**p=0.050** | **r=-0.348,**<br>**p<0.001** | **r=-0.288,**<br>**p=0.013** |
| **Monocyte,**<br>**%** | r=-0.116,<br>p=0.167 | r=-0.044,<br>p=0.601 | **r=-0.243,**<br>**p=0.003** | r=-0.194,<br>p=0.097 |

LDL-C, low-density lipoprotein cholesterol; HDL-C, high-density lipoprotein cholesterol; TG, triglycerides; apoB, apolipoprotein B; Lp(a), lipoprotein (a). Significant Spearman's correlations and p-values are in bold.

**Table 16.** Correlations between lipid and calculated total blood count readings.

| **Readings** | **Total**<br>**cholesterol**<br>**concentration** | **LDL** | **HDL** | **TG** | **ApoB** | **Lp(a)** |
| --- | --- | --- | --- | --- | --- | --- |
| **PLR** | r=0.014,<br>p=0.867 | r=0.024,<br>p=0.771 | **r=0.176,**<br>**p=0.035** | r=-0.051,<br>p=0.554 | r=0.040,<br>p=0.741 | r=0.206,<br>p=0.079 |
| **LMR** | **r=0.220,**<br>**p=0.008** | r=0.156,<br>p=0.059 | **r=0.264,**<br>**p=0.001** | r=0.025,<br>p=0.775 | r=0.222,<br>p=0.061 | r=0.170,<br>p=0.147 |
| **MLR** | **r=-0.221,**<br>**p=0.008** | r=-0.157,<br>p=0.058 | **r=-0.265,**<br>**p=0.001** | r=-0.025,<br>p=0.774 | r=-0.222,<br>p=0.060 | r=-0.171,<br>p=0.146 |
| **SIRI** | **r=-0.206,**<br>**p=0.013** | r=-0.159,<br>p=0.054 | **r=-0.262,**<br>**p=0.002** | r=0.051,<br>p=0.548 | **r=-0.261,**<br>**p=0.027** | r=-0.158,<br>p=0.179 |
| **MHR** | **r=-0.353,**<br>**p<0.001** | **r=-0.245,**<br>**p=0.003** | **r=-0.801,**<br>**p<0.001** | **r=0.170,**<br>**p=0.046** | r=-0.209,<br>p=0.083 | **r=-0.364,**<br>**p=0.002** |

LDL-C, low-density lipoprotein cholesterol; HDL-C, high-density lipoprotein cholesterol; TG, triglycerides; apoB, apolipoprotein B; Lp(a), lipoprotein (a); PLR, platelet to lymphocyte ratio; LMR, lymphocyte to monocyte ratio; MLR, monocyte to lymphocyte ratio; SIRI, systemic inflammation response index; MHR, monocyte-to-high-density- lipoprotein ratio. Significant Spearman's correlations and p-values are in bold.

NMR very weakly correlated with HDL *(r=0.184, p=0.028).* No significant correlations were found between NLR, SII and lipidogram readings.

PLR values weakly correlated with leukocyte counts *(r=-0.362, p<0.001)* and NLR, NMR, SII values *(r=0.284, p<0.001; r=0.256, p<0.001; r=0.295, p<0.001,* respectively) and moderately correlated with SIRI and MHR values *(r=0.532, p<0.001; r=0.542, p<0.001,* respectively).

There was a weak correlation between a higher platelet count and lymphocyte count *(r=0.220, p=0.005),* a percentage of monocytes *(r=-0.263, p<0.001)* and a very weak correlation with monocyte count *(r=-0.164, p=0.035).* Platelet counts weakly correlated with PLR, NMR and LMR values *(r=0.398, p<0.001; r=0.208, p=0.007; r=0.301, p<0.001,* respectively) and with MLR *(r= −0.301, p<0.001)* and MHR *(r=-0.231, p=0.005).* SIRI correlated very weakly with platelet count *(r=-0.170, p=0.029).* In contrast, MPV correlated very weakly with platelet count and SII *(r= −0.197, p=0.011; r=-0.165, p=0.034,* respectively) and weakly with PLR *(r=-0.261, p<0.001)*.

#### 4.3 Correlation between calculated total blood count readings and severity of CAD

In terms of correlation with the severity of CAD, there was a positive mean correlation between SII and the severity of coronary stenosis according to the Gensini score *(r=0.511, p<0.001)* (Figure 3).

**Figure 3.**
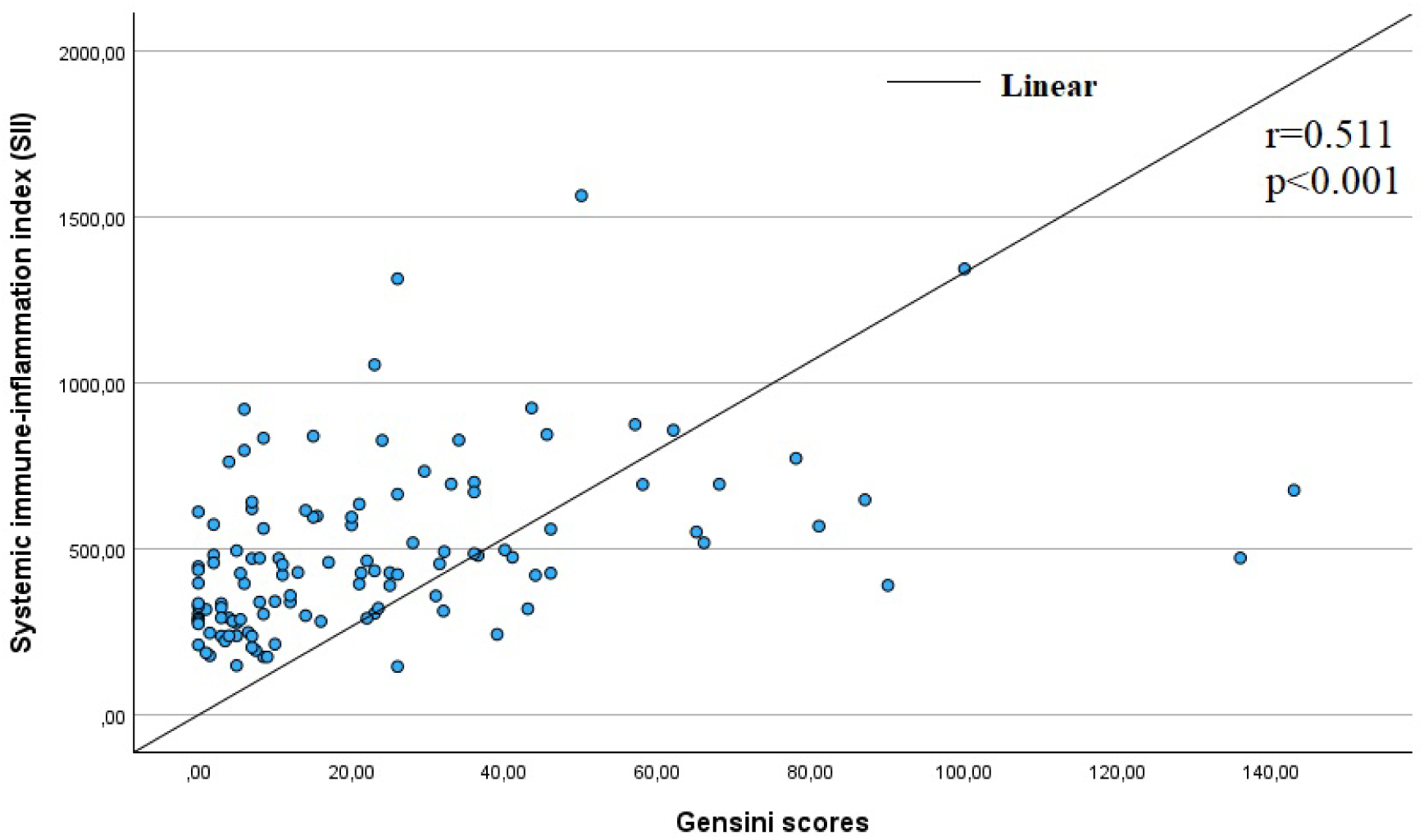
Correlation of SII with the degree of coronary artery disease according to the Gensini score.

The severity of coronary artery stenosis as assessed by the Gensini score was found to be weakly correlated with PLR, NMR, MLR *(r=0.265, p=0.004; r=0.269, p=0.003; r=0.356, p<0.001,* respectively) and LMR *(r=-0.355, p<0.001)* and moderately correlated with NLR and SIRI *(r=0.567, p<0.001; r=0.474, p<0.001,* respectively).

In addition, the CAD-RADS classification showed that the severity of coronary stenosis was weakly correlated with NLR, SII and MHR *(r=0.365, p<0.001; r=0.239, p=0.018; r=0.346, p<0.001*, respectively) and moderately correlated with LMR *(r=-0.454, p<0.001),* MLR and SIRI *(r=0.455, p<0.001; r=0.522, p<0.001,* respectively). Clearly, there was a statistically significant strong positive correlation between CAD-RADS and Gensini score *(r=0.674, p<0.001)*.

#### 4.4. Correlations related to readings of baseline characteristics

##### 4.4.1. Correlations between baseline and lipid readings

Our study showed that smoking was weakly but statistically significantly associated with levels of triglycerides *(r=0.357, p=0.003)* and levels of HDL-C *(r= −0.252, p=0.034).* BMI correlated with HDL-C levels *(r= −0.299, p=0.04)*. Older age correlated with dyslipidaemia *(r=0.238, p=0.026).* Triglyceride levels weakly correlated with LVEDDi *(r= −0.275, p=0.033),* e′_sep_ *(r= −0.399, p=0.014),* and e′_lat_*(r= −0.341, p=0.039).* Uric acid levels correlated with HDL-C *(r= −0.339, p=0.030)* and triglycerides levels *(r=0.336, p=0.037)*.

##### 4.4.2. Correlation between baseline and echocardiography readings

Age correlated with LVEF *(r= −0.239, p=0.028)* and PWT *(r= −0.262, p=0.021);* diastolic blood pressure correlated with PWT *(r= 0.268, p=0.018),* LVM *(r= 0.306, p=0.007)* and RV S’ *(r= 0.302, p=0.011).* BMI correlated with LVEDD *(r= 0.340, p=0.003).* Presence of AH correlated with IVS thickness *(r= 0.242, p=0.034)*, LVM *(r= 0.316, p=0.005)* and LVMi *(r= 0.331, p=0.003). S*moking weakly correlated with LVEDDi *(r=0.357, p=0.002).* In addition, e′_sep_ moderately correlated with uric acid levels *(r= −0.429, p=0.036)*.

##### 4.4.3. Correlations between baseline and total blood count and inflammation readings

Smoking correlated with uric acid concentration (*r=0.295, p=0.044) and erythrocyte count (r=0.371, p=0.001).* AH correlated with leukocyte, neutrophil and monocyte counts (Table 17). Obesity correlated with uric acid levels *(r=0.299, 0.041). A*n early history of cardiovascular disease correlated with neutrophil percentage *(r=0.230, p=0.036)*. Lymphocyte counts correlated with systolic and diastolic blood pressure (*r=0.223, p=0.041; r=0.280, p=0.01,* respectively).

**Table 17.**
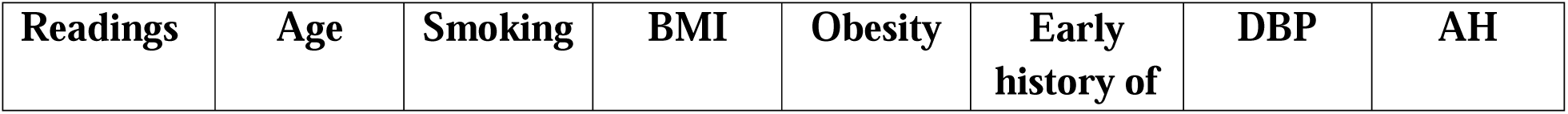
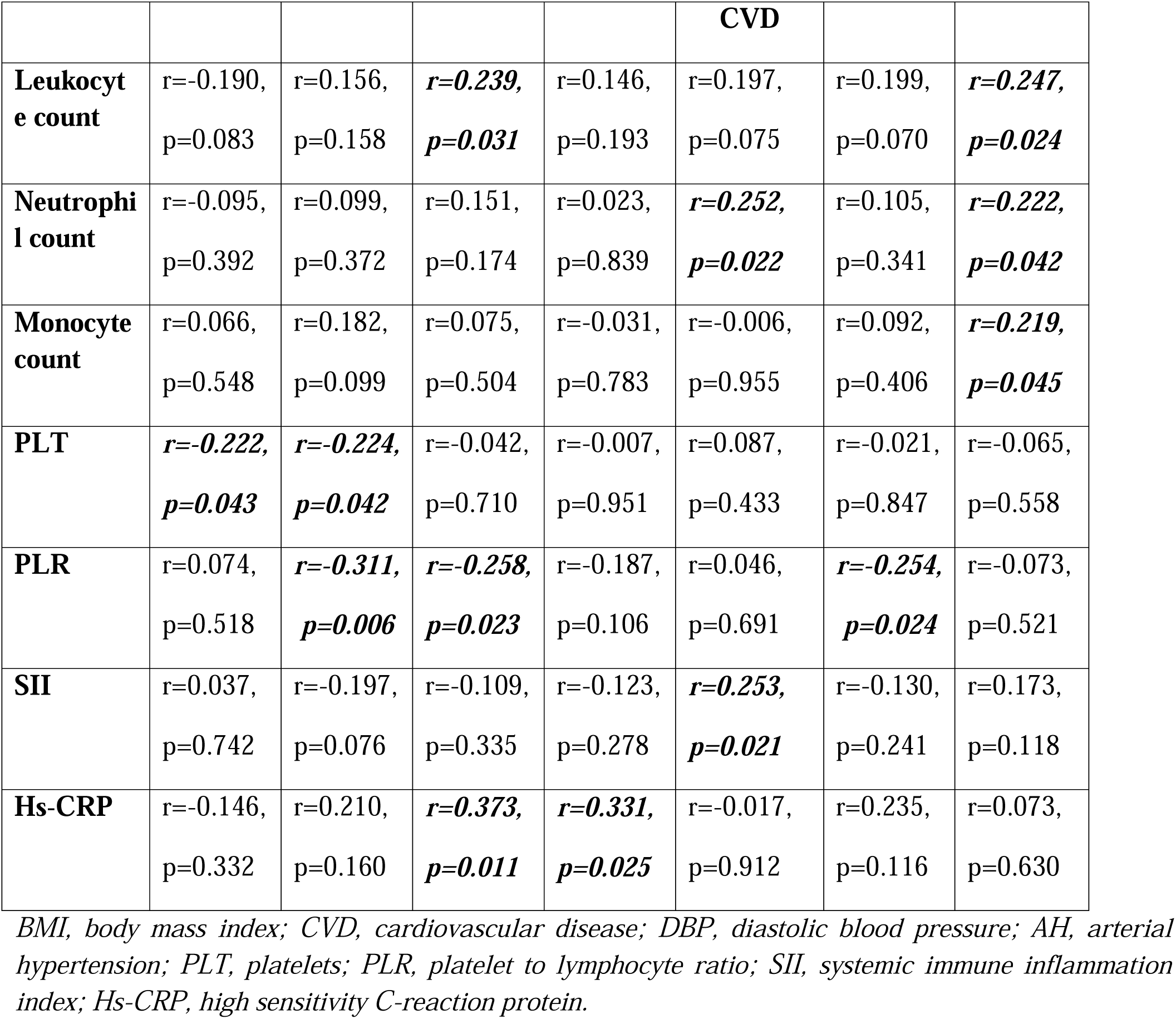
Correlations between baseline and total blood count and inflammation readings.

| Readings | Age | Smoking | BMI | Obesity | Early history of | DBP | AH |
| --- | --- | --- | --- | --- | --- | --- | --- |

|  |  |  |  |  | **CVD** |  |  |
| --- | --- | --- | --- | --- | --- | --- | --- |
| **Leukocyte count** | r=-0.190,<br>p=0.083 | r=0.156,<br>p=0.158 | ***r=0.239,***<br>***p=0.031*** | r=0.146,<br>p=0.193 | r=0.197,<br>p=0.075 | r=0.199,<br>p=0.070 | ***r=0.247,***<br>***p=0.024*** |
| **Neutrophil count** | r=-0.095,<br>p=0.392 | r=0.099,<br>p=0.372 | r=0.151,<br>p=0.174 | r=0.023,<br>p=0.839 | ***r=0.252,***<br>***p=0.022*** | r=0.105,<br>p=0.341 | ***r=0.222,***<br>***p=0.042*** |
| **Monocyte count** | r=0.066,<br>p=0.548 | r=0.182,<br>p=0.099 | r=0.075,<br>p=0.504 | r=-0.031,<br>p=0.783 | r=-0.006,<br>p=0.955 | r=0.092,<br>p=0.406 | ***r=0.219,***<br>***p=0.045*** |
| **PLT** | ***r=-0.222,***<br>***p=0.043*** | ***r=-0.224,***<br>***p=0.042*** | r=-0.042,<br>p=0.710 | r=-0.007,<br>p=0.951 | r=0.087,<br>p=0.433 | r=-0.021,<br>p=0.847 | r=-0.065,<br>p=0.558 |
| **PLR** | r=0.074,<br>p=0.518 | ***r=-0.311,***<br>***p=0.006*** | ***r=-0.258,***<br>***p=0.023*** | r=-0.187,<br>p=0.106 | r=0.046,<br>p=0.691 | ***r=-0.254,***<br>***p=0.024*** | r=-0.073,<br>p=0.521 |
| **SII** | r=0.037,<br>p=0.742 | r=-0.197,<br>p=0.076 | r=-0.109,<br>p=0.335 | r=-0.123,<br>p=0.278 | ***r=0.253,***<br>***p=0.021*** | r=-0.130,<br>p=0.241 | r=0.173,<br>p=0.118 |
| **Hs-CRP** | r=-0.146,<br>p=0.332 | r=0.210,<br>p=0.160 | ***r=0.373,***<br>***p=0.011*** | ***r=0.331,***<br>***p=0.025*** | r=-0.017,<br>p=0.912 | r=0.235,<br>p=0.116 | r=0.073,<br>p=0.630 |

*BMI, body mass index; CVD, cardiovascular disease; DBP, diastolic blood pressure; AH, arterial hypertension; PLT, platelets; PLR, platelet to lymphocyte ratio; SII, systemic immune inflammation index; Hs-CRP, high sensitivity C-reaction protein.*

Age weakly correlated with lower erythrocyte count *(r= −0.324, p=0.003)*, lymphocyte count *(r= −0.291, p=0.034),* and diastolic BP *(r= −0.365, p<0.001).* There were found some weak correlations between calculated total blood count and baseline readings (Table 17).

#### 4.5. Correlations related to drug usage

Some weak correlations have been found between the readings of lipid concentration, total blood counts and drug use (Tables 18 and 19). Additional correlations were found: the use of beta-blockers, ACEi or ARB and statin was associated with age *(r= 0.301, p<0.001; r= 0.374, p<0.001; r=0.198, p=0.010*, respectively); leukocyte counts correlated with statin *(r= 0.157, p=0.044)* and aspirin *(r= 0.223, p=0.004)* use, while platelet counts correlated negatively with statin use *(r= −0.211, p=0.007).* There were no significant associations between aspirin and CaCB and lipid and uric acid readings. Total blood readings were not associated with CaCB and MRA.

**Table 18.**
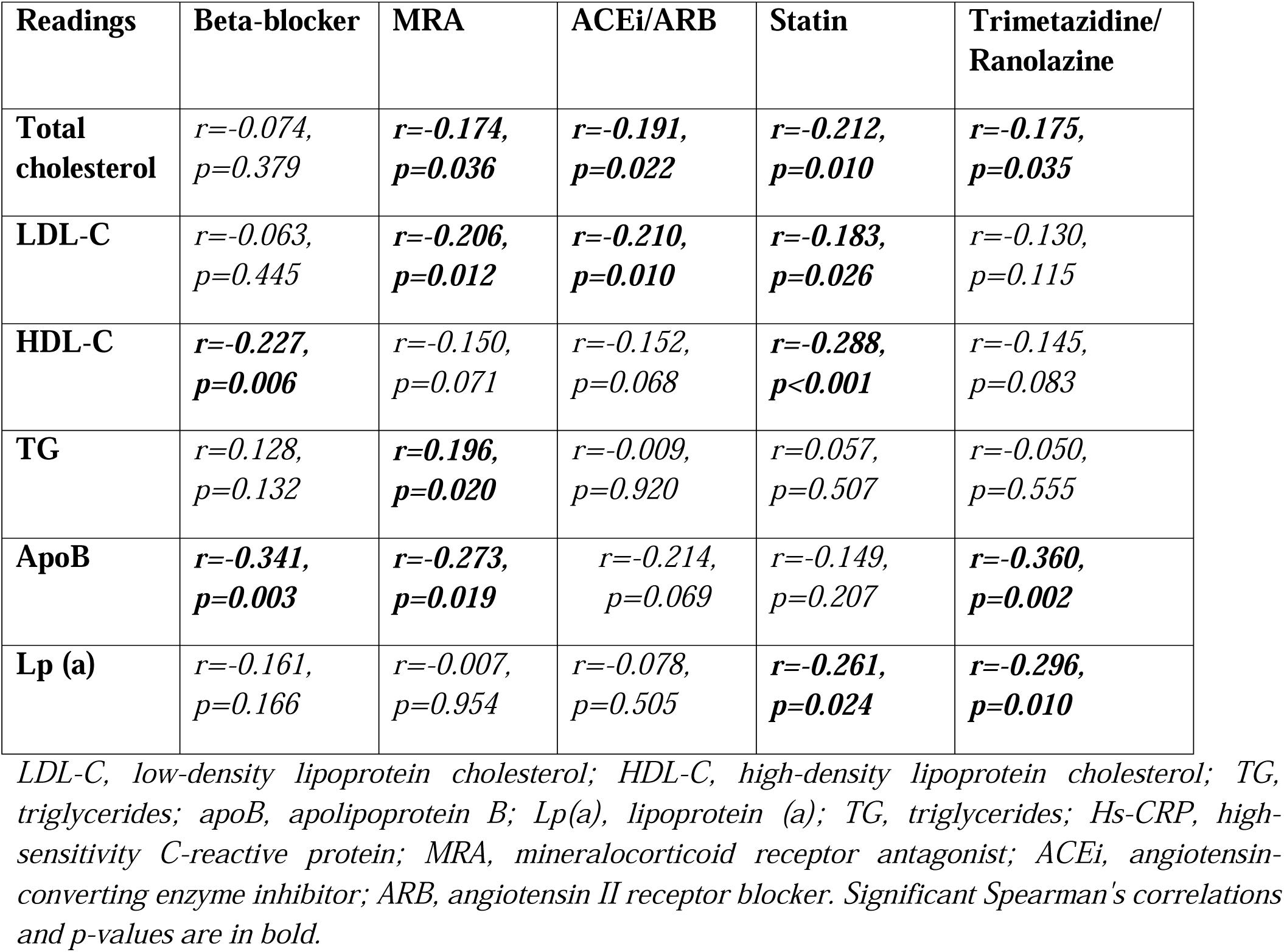
Drug usage correlations with lipid and uric acid readings.

| Readings | Beta-blocker | MRA | ACEi/ARB | Statin | Trimetazidine/<br>Ranolazine |
| --- | --- | --- | --- | --- | --- |
| **Total cholesterol** | $r=-0.074$,<br>$p=0.379$ | $r=-0.174$,<br>$p=0.036$ | $r=-0.191$,<br>$p=0.022$ | $r=-0.212$,<br>$p=0.010$ | $r=-0.175$,<br>$p=0.035$ |
| **LDL-C** | $r=-0.063$,<br>$p=0.445$ | $r=-0.206$,<br>$p=0.012$ | $r=-0.210$,<br>$p=0.010$ | $r=-0.183$,<br>$p=0.026$ | $r=-0.130$,<br>$p=0.115$ |
| **HDL-C** | $r=-0.227$,<br>$p=0.006$ | $r=-0.150$,<br>$p=0.071$ | $r=-0.152$,<br>$p=0.068$ | $r=-0.288$,<br>$p<0.001$ | $r=-0.145$,<br>$p=0.083$ |
| **TG** | $r=0.128$,<br>$p=0.132$ | $r=0.196$,<br>$p=0.020$ | $r=-0.009$,<br>$p=0.920$ | $r=0.057$,<br>$p=0.507$ | $r=-0.050$,<br>$p=0.555$ |
| **ApoB** | $r=-0.341$,<br>$p=0.003$ | $r=-0.273$,<br>$p=0.019$ | $r=-0.214$,<br>$p=0.069$ | $r=-0.149$,<br>$p=0.207$ | $r=-0.360$,<br>$p=0.002$ |
| **Lp (a)** | $r=-0.161$,<br>$p=0.166$ | $r=-0.007$,<br>$p=0.954$ | $r=-0.078$,<br>$p=0.505$ | $r=-0.261$,<br>$p=0.024$ | $r=-0.296$,<br>$p=0.010$ |

LDL-C, low-density lipoprotein cholesterol; HDL-C, high-density lipoprotein cholesterol; TG, triglycerides; apoB, apolipoprotein B; Lp(a), lipoprotein (a); TG, triglycerides; Hs-CRP, high-sensitivity C-reactive protein; MRA, mineralocorticoid receptor antagonist; ACEi, angiotensin-converting enzyme inhibitor; ARB, angiotensin II receptor blocker. Significant Spearman's correlations and p-values are in bold.

**Table 19.** Correlations between the total blood readings and drug usage.

| Readings | Beta-blocker | ACEi/ARB | Statin | Aspirin |
| --- | --- | --- | --- | --- |
| **Lymphocyte, %** | $r=-0.186$,<br>$p=0.016$ | $r=-0.213$,<br>$p=0.006$ | $r=-0.267$,<br>$p<0.001$ | $r=-0.282$,<br>$p<0.001$ |
| **Neutrophil count** | $r=0.139$,<br>$p=0.075$ | $r=0.176$,<br>$p=0.024$ | $r=0.194$,<br>$p=0.012$ | $r=0.247$,<br>$p=0.002$ |
| **Neutrophil, %** | $r=0.145$,<br>$p=0.064$ | $r=0.173$,<br>$p=0.026$ | $r=0.202$,<br>$p=0.009$ | $r=0.205$,<br>$p=0.009$ |
| **NLR** | $r=0.176$,<br>$p=0.024$ | $r=0.203$,<br>$p=0.009$ | $r=0.253$,<br>$p=0.001$ | $r=0.261$,<br>$p<0.001$ |
| **LMR** | ***$r=-0.157$,***<br>***$p=0.044$*** | ***$r=-0.162$,***<br>***$p=0.038$*** | ***$r=-0.244$,***<br>***$p=0.002$*** | ***$r=-0.183$,***<br>***$p=0.020$*** |
| **MLR** | ***$r=0.157$,***<br>***$p=0.044$*** | ***$r=0.161$,***<br>***$p=0.039$*** | ***$r=0.243$,***<br>***$p=0.002$*** | ***$r=0.183$,***<br>***$p=0.020$*** |
| **SII** | *$r=0.088$,*<br>*$p=0.262$* | *$r=0.152$,*<br>*$p=0.051$* | *$r=0.126$,*<br>*$p=0.108$* | ***$r=0.254$,***<br>***$p=0.001$*** |
| **SIRI** | ***$r=0.188$,***<br>***$p=0.016$*** | ***$r=0.218$,***<br>***$p=0.005$*** | ***$r=0.248$,***<br>***$p=0.001$*** | ***$r=0.258$,***<br>***$p<0.001$*** |
| **MHR** | ***$r=0.183$,***<br>***$p=0.028$*** | *$r=0.121$,*<br>*$p=0.148$* | ***$r=0.246$,***<br>***$p=0.003$*** | ***$r=0.175$,***<br>***$p=0.038$*** |

*ACEi, angiotensin-converting enzyme inhibitor; ARB, angiotensin II receptor blocker; NLR, neutrophil to lymphocyte ratio; LMR, lymphocyte to monocyte ratio; MLR, monocyte to lymphocyte ratio; SII, systemic immune-inflammation index; SIRI, systemic inflammation response index; MHR, monocyte-to-high-density- lipoprotein ratio. Significant Spearman's correlations and p-values are in bold.*

**Table 20.**
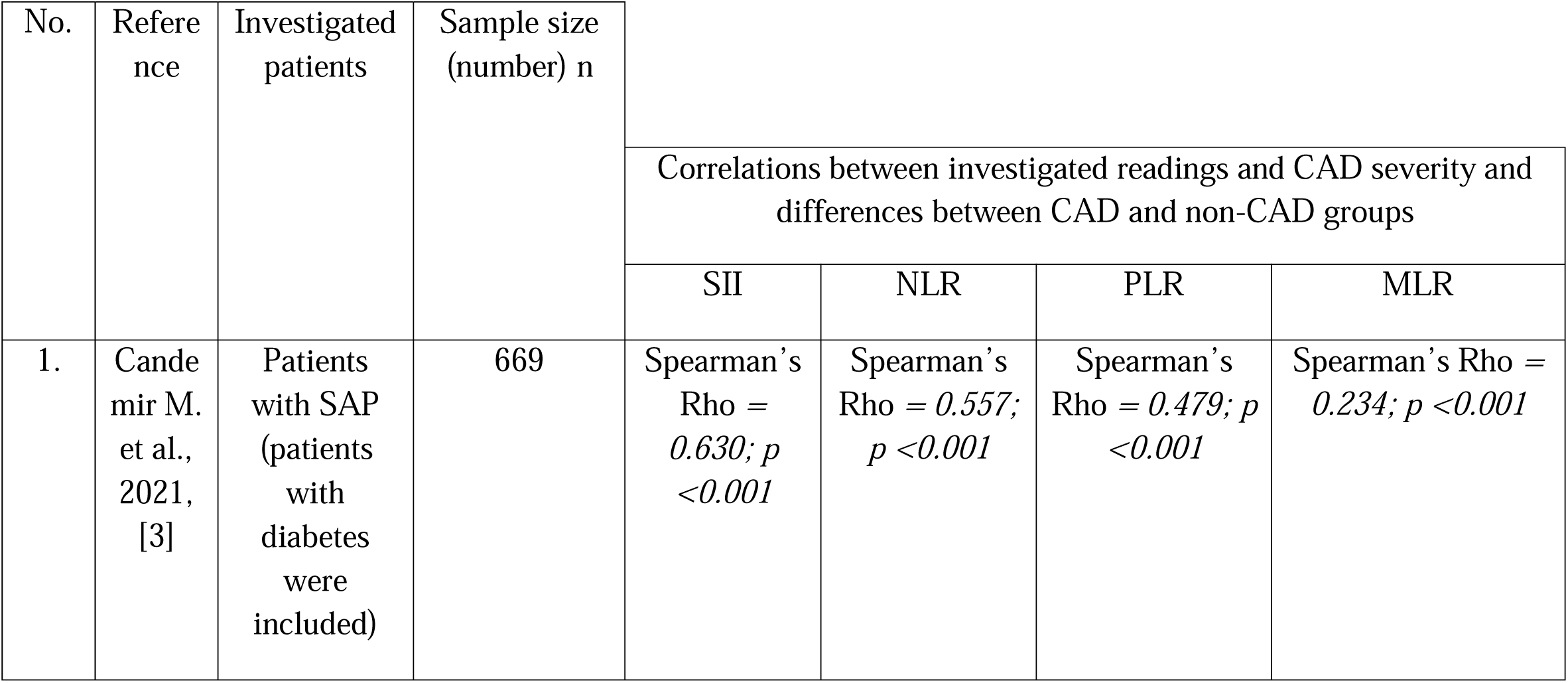
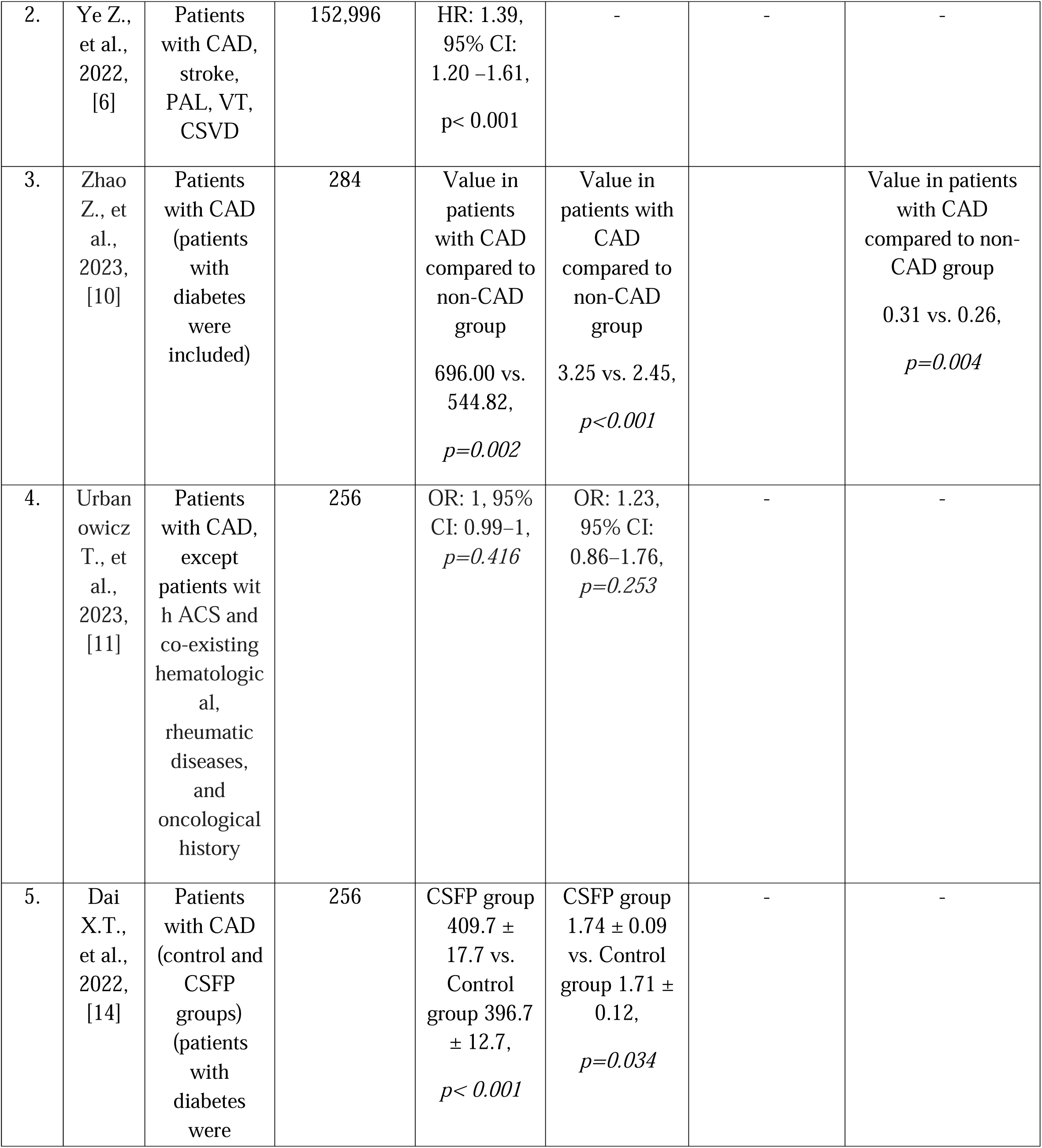
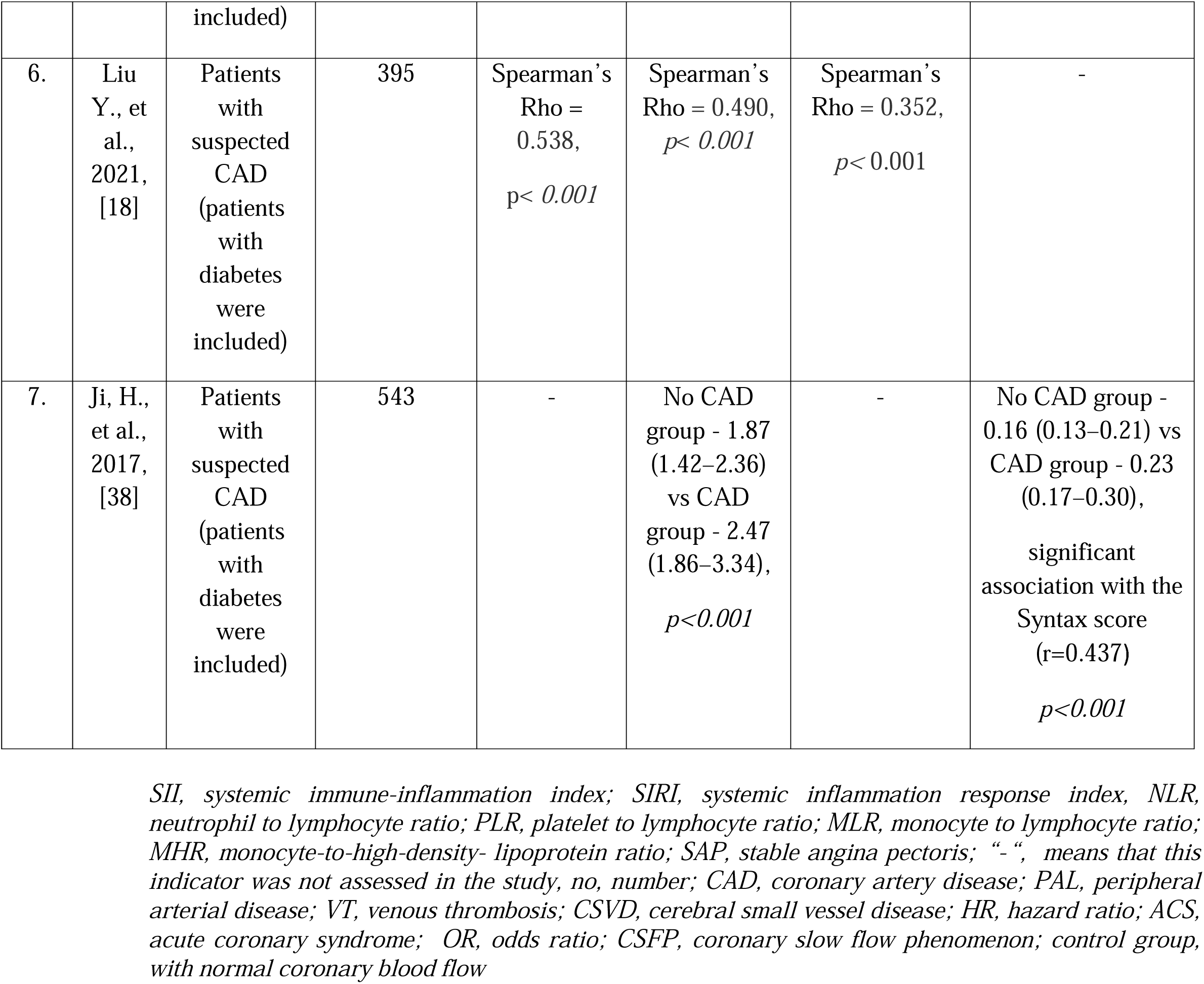
Literature findings of the relationship between calculated total blood count readings and the severity of coronary atherosclerosis and presence of CAD.

### 5. Discussion

This pilot retrospective study of patients with SAP shows that an easily measurable haematological markers, such as SII, SIRI, NLR, MHR, MLR, and LMR could indicate the severity of atherosclerosis. LMR and SIRI seems could be the best readings for chronic inflammation evaluation in the SAP patients. SIRI seems could reflects the degree of atherosclerosis best of all other calculated total blood count readings.

#### 5.1. Differences in laboratory parameters (calculated total blood counts readings, lipids and hs-CRP) between the patients’ groups

Our study showed that patients with CAD had higher NLR, SIRI, MLR, SII and lower LMR in comparison to patients without CAD. This can be explained by pathogenetic mechanisms, which have been described in detail in some publications.

For instance, neutrophils secrete neutrophil extracellular traps - pro-inflammatory and pro-thrombotic substances that trap leukocytes and promote thrombosis [5, 6, 32]. In patients with severe CAD, neutrophil counts are often elevated, primarily due to acute and chronic inflammatory reactions, tissue repair mechanisms and stress-induced mobilisation [6]. In addition, neutrophils can infiltrate endothelial tissue and release pro-oxidants and pro-inflammatory mediators, which in turn can form neutrophil extracellular traps and promote the formation and development of atherosclerotic plaques [44]. It should be mentioned that recent studies have shown that neutrophils play an important role in the production of reactive oxygen species and the environment during thrombus formation [13, 22]. In our patients neutrophil count was significantly higher in CAD group (Table 7) and increased with increasing in Gensini (Table 9) and CAD-RADS score number (Table 11). Reduced lymphocyte numbers could associate with the progression of atherosclerosis [5, 6, 33]. Our results shew lower lymphocyte count in patients with CAD (Table 7) and gradually decreasing with increasing degree of atherosclerosis (Tables 9 and 11). The combination of neutrophil and lymphocyte parameters are presented as better prognostic value than either of them alone [32, 33]. In our study, NLR was found higher in CAD group (Table 8) and increased in Gensini (Table 10) and CAD-RADS groups too (Table 12). Even more, the results of a study conducted on NLR in patients with CAD recently showed that increased NLR was a predictor of CAD prognosis (by a factor of 2.79) [34]. Thus, our study was in consensus with these results, as we also found that patients with CAD had a significantly higher NLR compared with patients without CAD. In addition, it seems NLR could be a useful marker for evaluating the severity of CAD.

Some studies have shown that high monocyte and low lymphocyte counts are independent predictors of CAD risk, and the MLR index integrates the risk of coronary lesion severity [35–38]. As mentioned earlier, it has been hypothesised that monocytes may enter the arterial wall, differentiate into macrophages and activate the secretion of pro-inflammatory cytokines, the production of matrix metalloproteinases and the production of reactive oxidative species, which are key players in the initiation of atherosclerotic plaque initiation and the formation or rupture of atherosclerotic plaques [5, 6, 36]. Lymphocytes are potentially important immune cells in cardiovascular disease, and low lymphocyte numbers have been shown to reflect impaired coronary microcirculation, which has been confirmed as an important mechanism in the pathogenesis of CAD [5, 6, 37]. We found that MLR was significantly higher in patients with CAD (Table 8). Ji H et al. reported similar results in their study and confirmed that MLR may be a risk factor for atherosclerosis and correlate with CAD severity. Their results showed that MLR > 0.18 predicted CAD with a sensitivity of 69.03% and specificity of 64.81% [38]. It should be mentioned that we not found the difference in monocyte count between the groups according to CAD presence (Table 7) and Gensini score (Table 9). But monocyte count increases gradually when increased CAD-RADS group number (Table 11). Our results shew MLR could give more information related to patients’ condition related to atherosclerosis as cell count.

Kose N. et al. attempted to assess the association between LMR and CAD in patients with stable angina pectoris in their study. Their exclusion criteria were similar to ours. The results of their study showed that LMR was an independent predictor of CAD severity in patients with stable CAD. Their results showed that patients with CAD had lower LMR values (4.5±3.2 vs. 6±2.9, p<0.001) compared to the group without CAD [39]. In our study, the same trend of LMR values was also observed in these groups (Table 8). Recent studies show that LMR can also be used as a marker of inflammation, just like the other markers mentioned above - NLR, MLR. The LMR combines two independent markers of inflammation, and both high monocyte counts and low lymphocyte counts are associated with coronary atherosclerosis [5, 6, 36, 37, 40]. Monocytes are recruited in the intima and subintima, differentiate into macrophages and mast cells in response to a number of locally produced cytokines, and initiate the atherosclerotic process by supporting plaque formation [41]. Meanwhile, lymphocytes play an important role in the pathogenesis of atherosclerosis by modulating the immune response, with different subtypes promoting or inhibiting plaque growth and stability. Lymphocytes, especially T cells, play a key role in modulating the immune response in atherosclerotic plaques. Regulatory T cells (Tregs) have been shown to have anti-inflammatory properties and attenuate atherosclerosis by reducing inflammation in atherosclerotic plaques [42]. The combined role of lymphocytes and monocytes in atherosclerosis highlights the chronic inflammatory process of the disease and may lead to the progression of atherosclerosis [40–42]. So our found decreased LMR, increased MLR in patients with CAD and decreasing LMR and increasing MLR with increasing Gensini score and CAD-RADS group number supplements recent research findings.

In addition, other indicators calculated from a complete blood count may be relevant to the state of inflammation and the pathogenesis of atherosclerosis too. Urbanowicz T. and co-authors attempted to assess the predictive role of SIRI for CAD in their retrospective study. Their study included 256 patients and excluded patients with acute coronary syndromes or comorbid haematological, rheumatic and oncological diseases. The results of their study were similar to our study and confirmed the trend that SIRI was the highest in patients with more severe CAD (complex (2- or 3-vessel) coronary disease). For example, in their study, the median SIRI was the highest in the group of patients with complex CAD - 0.99 (0.76-1.27), compared with the group without CAD - 0.82 (0.57-1.06), p<0.001. In addition, a logistic regression analysis performed in their study in patients with single-vessel CAD compared to patients without coronary disease showed that SIRI can be a predictive laboratory parameter (OR: 3.32, 95% CI: 1.56–7.03, p=0.002)[11]. As well and our results showed that patients who underwent CA and their CAD severity was assessed by the Gensini score also had the highest SIRI in the group with the most severe CAD (Group 3, Table 10). The same trend was observed in our patients who underwent CCTA and coronary disease severity was assessed by the CAD-RADS score (Group 4, Table 12). It seems that elevated SIRI levels, which indicate an imbalance between high neutrophil and monocyte counts and low lymphocyte counts, could be indicative of an increased pro-inflammatory state. A high SIRI value could be a strong indication of increased risk of atherosclerosis, as it indicates an overactive immune response (involving even three types of leukocytes) that contributes to the formation and progression of atherosclerotic plaques. It is thought that low SIRI values may indicate a more balanced immune response and better regulation of inflammation, which may protect against atherosclerosis [11, 43].

Two years ago, Ye Z. et al. published a systematic review supporting the idea that SII may be another prognostic marker of atherosclerosis, which, compared to traditional markers of inflammation, may be a valuable tool for predicting CAD risk, as it reflects the balance between inflammation and immunity, which is of great importance in the CAD context. High SII may be associated with increased inflammatory activity due to high neutrophil and platelet counts and low lymphocyte concentrations [6]. Scientists are still trying to find the answer to why SII increases in CAD. It is thought that in CAD, platelets are activated and accumulate at sites of vascular damage, contributing to thrombosis and subsequent inflammation. Thus, increased platelet activity is associated with systemic inflammation and may lead to higher SII values [14]. Additionally, leukocytes and monocytes, recruited by platelets, migrate to the site of inflammation and release inflammatory mediators such as chemokines and cytokines, which can trigger vascular inflammation. As already mentioned, monocytes play a key role in this inflammatory process by migrating to the site of atherosclerotic lesions, where they differentiate into macrophages and contribute to plaque formation. Thus, patients with severe CAD usually have elevated monocyte counts, mainly due to chronic inflammation, endothelial dysfunction and elevated cytokine levels [14]. On the other hand, the decrease in lymphocyte counts in patients with severe CAD is primarily the result of chronic inflammation, dysregulation of the immune system, acute stress response and possible migration of lymphocytes to inflamed tissues. In addition, the pro-inflammatory environment often leads to apoptosis of lymphocytes, resulting in a reduction in their number [44]. Our data showed that SII was higher in the CAD group in comparison with group without CAD. Accordingly, SII gradually increased with increasing in Gensini score group. But it not differ between the CAD-RADS score groups.

Hs-CRP is a valuable inflammatory factor in coronary disease as it is a marker of systemic inflammation, which plays an important role in the pathophysiology of atherosclerosis. Elevated levels of hs-CRP reflect a state of inflammation that can be related with endothelial dysfunction, plaque formation and to be associated with increased plaque instability and vulnerability [45]. This is the reason, why we chose to evaluate the hs-CRP in our study.

Behera D. et al. confirmed the importance of hs-CRP as a significant risk factor for CAD. The results showed that hs-CRP levels were significantly higher in the group of patients with CAD compared with patients without CAD (2.932 ± 0.605 vs. 0.379 ± 0.202 mg/dl, p < 0.001) [46]. Their results were similar to ours, and we also found significantly higher hs-CRP values in the CAD group compared to the group without CAD (Table 6).

Cederström S. et al. recently analysed the relationship between hs-CRP and the degree of atherosclerosis in CAD. Their population cohort was large, comprising 25 408 patients, but the results showed that elevated hs-CRP may be only weakly associated with the presence of coronary atherosclerosis [19]. Also, we found no significant changes in hs-CRP levels when assessing the severity of CAD. Similar results were reported by Bouzidi N et al. in their study, who also concluded that hs-CRP levels were not associated with the severity of CAD, as assessed by the degree of stenosis and the number of coronary vessels affected [47].

We found that lipids concentration not differ between all the investigated groups. Other research work founsimilarly: no statistically significant differences between total cholesterol, triglycerides, LDL-C and uric acid levels in the two groups of patients with and without CAD [18]. We think the absence of differences could be related to statins usage.

In conclusion it could be stated that our results showed NLR, MLR, SIRI and SII were higher and LMR was lower in CAD group. NLR, MLR, SIRI and SII increased and LMR decreased gradually with increasing of Gensini score group. There is the tendency MHR and SIRI to increase and LMR to decrease when the group number of CAD-RADS increase. SII not differ between the CAD-RADS groups. LMR and SIRI increased gradually with increasing of Gensini score group. So, LMR and SIRI seems could be the best readings for chronic inflammation evaluation in patients with SAP, especially when hs-CRP not differ between the investigated groups according to Gensini and CAD-RADS score.

#### 5.2. Discussion of correlations

##### 5.2.1. Correlation between readings based on total blood count data and lipids and hs-CRP

Our study showed that lipidogram reading HDL-C level reversibly correlated with leukocyte, neutrophil and monocyte counts and percentage of monocytes and MLR, SIRI and MHR, while HDL-C levels were correlated only weakly with PLR and LMR. But neither triglyceride, nor HDL-C differ between the patients grouped according to CAD presence, Gensini score and CAD-RADS classification in our study (Tables 7-9). So, it seems only HDL-C concentration could be related with leukocyte, monocyte, and neutrophil counts in SAP patients.

It is known that elevated lipid levels can lead to increased involvement and activation of immune cells, which promote the progression of CAD and associated with cardiovascular risk. When lipids, especially oxidised LDL, accumulate in the intima of the artery, they activate the immune system, leading to an increase in the production of leucocytes and the recruitment of monocytes and neutrophils to the site of inflammation. Lipid concentration is presented as closely linked to the number of leukocytes, monocytes and neutrophils through a variety of mechanisms related to inflammation, immune activation and atherosclerotic processes [48]. One study showed that levels of HDL-C associated reversibly with MLR and SIRI, mainly due to HDL-C’s anti-inflammatory properties. In addition, HDL-C may inhibit the expression of endothelial cell adhesion molecules, thereby reducing the recruitment of immune cells such as monocytes and lymphocytes to inflamed tissues. Higher levels of HDL-C may stimulate the differentiation of T regulatory cells, which may help to suppress inflammatory reactions and reduce lymphocyte activation, thus contributing to lower MLR. In addition, the ability of HDL-C to transport cholesterol, reduce inflammation, improve endothelial function and modulate immune responses contributes to the inverse relationship between HDL-C levels and SIRI [49]. Our findings are in concensus with these findings. In our study HDL-C reversibly correlated with MLR and SIRI and positively correlated with LMR, despite the fact that lipid concentration not differ between all the groups analysed.

As already mentioned, monocytes are involved in atherosclerosis by migrating into the subendothelial space and taking up lipoproteins. In contrast, HDL-C has anti-atherosclerotic properties and HDL-C molecules prevent monocyte activation and recruitment [10]. So, it can be assumed that higher HDL-C levels (reflected by lower MHR) reflect an increased cholesterol efflux capacity leading to reduced lipid accumulation in macrophages and reduced mast cell formation, as found in our study. We found that MHR correlated with all the lipid readings (total cholesterol, LDL, HDL and Lp(a) concntrations) except the apoB (Table 15). Thus, our results supplement findings about the relationship between lipid levels and inflammatory statement in the atherosclerosis by possibly modulating inflammatory reactions.

Our study showed that hs-CRP levels positively correlated with neutrophil counts and percentage, NLR, NMR, SIRI and reversibly correlated with percentage of lymphocytes. He J. et co-authours also found that NLR levels were modestly, and significantly correlated with hs-CRP levels. This study demonstrated discordantly elevated NLR levels were associated with a greater risk of adverse clinical events in patients with stable CAD [50]. Bouzidi N. et al. in their study also found that inflammatory parameters - white blood cells were significantly increased in the patients with high hs-CRP levels as have shown our results too. In addition, increased hs-CRP levels often reflect ongoing inflammation, and neutrophils are one of the first immune cells to respond to inflammation [47]. So NLR, NMR, SIRI (readings that involve neutrophil count) correlations with hs-CRP are logical and could confirm low inflammation in SAP patients.

Concluding results about correlations between readings based on total blood count data and lipids and hs-CRP could be stated, that lipid concentration in the blood could be related with leukocyte, monocyte, lymphocyte and neutrophil counts in SAP patients. Hs-CRP correlation with NLR, NMR, SIRI confirm that NLR, NMR, SIRI shew the inflammation in these patients.

#### 5.2.1. Correlation between echocardiography readings and other estimated readings

Looking for correlation with cardiac echocardiography readings and laboratory parameters, we found that LVEF positively correlated with lymphocyte percentage and LMR, but negatively – with the count and percentage of neutrophil, NLR, and MLR. In addition, LVEDD correlated with erythrocyte count, neutrophil count and percentage, NLR, SIRI, MHR and reversibly - with lymphocyte percentage, platelet count, and LMR. LA diameter also negatively correlated with platelet count, lymphocyte percentage, LMR and positively - with MLR, SIRI and MHR. Hs-CRP directly correlated with IVS and LVPW thickness, LVM. Meanwhile, Medenwald D. and co-authors also found that hs-CRP, can be associated with structural changes in the heart, in particular the mass of the left ventricle and the thickness of the interventricular septum and the left ventricular posterior wall [26]. Previous studies have shown that markers of inflammation such as hs-CRB, high leukocyte counts and NLR values are associated with left ventricular diastolic dysfunction [27, 28]. Similarly, our study demonstrated that left ventricular diastolic function was correlated reversibly with count and percentage of lymphocytes, LMR and positively - with count of neutrophil, NLR, MLR and SIRI (Tables 13, 14). These relationships could show the asociations between the cardiac function and total blood count related inflammation readings.

##### 5.2.2. Correlation between readings based on total blood count data and severity of CAD

Several studies have looked at the relationship between calculated total blood count readings and the severity of coronary atherosclerosis (Table 16). Our study is unique in that we analysed all the possible readings on the basis of total blood count, compared to other published studies. According to our knowledge, we first evaluate calculated total blood count readings’ relationship with the degree of atherosclerosis in SAP patients without co-morbidities.

A growing number of studies have examined the role of SII in atherosclerosis and cardiovascular disease (CVD). In some research works it was showed that elevated SII levels associate with an increased risk of CAD and predict more severe CAD [3, 6]. Dai et al. estimated that elevated levels of SII significantly associate with the progression of atherosclerosis, which leads to slow coronary flow phenomenon, and associate with a higher number of diseased coronary vessels [14]. The recent meta-analysis mentioned above also showed that elevated SII levels are associated with an increased risk of CVD. In addition, some studies have hypothesised that systemic immune inflammation plays a key role in destabilising atherosclerotic plaques, leading to plaque rupture and subsequent development of acute coronary syndrome. Liu et al. also found a positive correlation between the SII and the severity of coronary artery disease. As in our study, they also showed that a higher SII correlated with more severe coronary atherosclerosis assessed by the Gensini score [18]. Gur et al. showed in their study that elevated SII was significantly associated with greater myocardial damage in acute coronary syndrome (ACS) [17]. In contrast to this study, in our study we excluded patients with ACS and patients with diabetes in order to have more homogeneous groups and to eliminate the effects of other conditions on the immune and inflammatory systems. Candemir M et al. showed that a SII value above 750 could predict severe CAD with 86.2% sensitivity and 87.3% specificity [3]. However, it is important to note that, although the results are encouraging, the level of evidence for the use of SII as a CAD biomarker remains generally low [6]. This could be due to a number of factors, including differences in study designs, patient cohorts and outcome measures across studies. Our results suggest that higher SII may reflect increased systemic inflammation, which in turn contributes to CAD progression. In our study, the SII positively correlated with Gensini (r=0.511, p<0.001) and CAD-RADS scores (r=0.239, p=0.018) (CAD severity) and could have a better predictive power for CAD onset than lipid fractions and hs-CRP.

Some authors looked for the other total blood count calculated readings. Candemir M et al. studied patients with SAP to assess the prediction of CAD severity by prognostic indicators derived from SII, NLR, PLR, MLR. Their results shew that NLR, PLR, MLR and SII statistically significantly correlated with CAD severity, but SII was suggested as a risk factor for atherosclerosis and as better predictor of CAD severity than other total blood count calculated readings such as NLR, MLR and PLR. [3].

Guo W. et al. found that patients with CAD had a higher NMR, and the increase in NMR was significantly associated with the severity of coronary atherosclerosis. The investigators of this study concluded that NMR may help to predict cardiovascular events in patients with CAD and may be associated with vulnerable plaque burden and a higher risk of adverse cardiovascular events [9]. Our results also showed that NMR was statistically significantly higher in patients with more severe CAD compared with patients without CAD, and the NMR value was significantly higher according to the Gensini score (Groups 2 and 3, Table 10). However, we found no significant differences and no correlation between NMR and CAD-RADS score. This could be explained by the comorbidities of patients in Guo W et al. groups.

Also, Liu Y. confirmed in his study that NLR can predict atherosclerosis with greater power, and NLR and PLR values positively correlated with atherosclerosis severity and Gensini score [18]. Other previous studies have also shown that NLR, PLR and MLR can predict the inflammatory role of different cell types in atherosclerosis and may be predictors of coronary morbidity and mortality in patients with CVD [23, 24, 25]. In addition, Núñez J. et al. also showed in their study that lower lymphocyte counts were associated with faster progression of atherosclerosis in coronary disease [20].

Meanwhile, Kiris T. et al. reported in their study that high MLR values were associated with vulnerable plaques in patients with stable angina [21]. These results also are in consensus with those of our study. We found that NLR, MLR, SIRI and SII were significant higher (Table 11, Figure 1) in patients with CAD. Furthermore, NLR, MLR, SII, SIRI and MHR were the highest and LMR – the lowest in Group 3 of patients grouped by Gensini score (Table 13). Accordingly, neutrophil perecentage was the lowest and lymphocyte percentage was the highest in the first Group of patients grouped by Gensini score (Table 12). Therefore, our results supplement knowledge that neutrophil, lymphocyte, and monocyte count and calculated total blood count readings NLR, MLR, LMR, SIRI, SII and MHR could be related with degree of atherosclerosis.

Last year, Chinese researchers Zhao et al. enrolled nearly 300 patients with suspected CAD confirmed by coronary angiography to assess prognostic indicators of systemic inflammation and factors related to lipid metabolism. Their study showed that MHR, NLR, MLR, NLR and SII levels were significantly higher in CAD patients than in non-CAD patients, and the results showed that an MHR value of more than 0.47 and an SII value of more than 589.12 were considered as independent CAD risk factors [10].

Yildiz et al. showed that the SII can help to assess not only the severity but also the prognosis of CAD. Higher values of the SII predicted a higher 1-year rate of major adverse cardiac events [30]. The SII reported to be a more accurate predictor of inflammatory activity compared to a single haematological parameter. The SII may serve as a biomarker to complement the prognostic information that traditional risk factors cannot provide [6, 30].

Several years ago, Chinese researchers Si Y. and colleagues organised a study to assess the relationship between LMR and coronary plaque burden associated with CAD. Their study population consisted of patients with suspected CAD with SAP symptoms. Exclusion criteria were acute coronary syndrome, renal failure, connective tissue diseases, severe valvular heart disease and hypertrophic cardiomyopathy, pregnancy, but diabetics were included. Coronary computer tomographic angiography was used to confirm coronary artery stenosis and coronary artery calcification (CAC). The results of this study showed that patients with CAD tended to have a lower LMR value than patients without CAD (p=0.001). Moreover, LMR was negatively correlated with CAC score and was an independent risk factor for CAC score (p < 0.05). The diagnostic cutoff point of LMR, calculated by multiple logistic regression model, showed that LMR ≤4.8 was a new independent risk factor for CAD (p < 0.05) [8]. Our results are in line with Si Y.’s findings: we found that LMR negatively correlated with coronary stenosis (CAD-RADS class number). Thus, it can be argued that low LMR, low lymphocyte counts, or high monocyte counts may induce inflammation and oxidative stress, releasing more pro-inflammatory factors, damaging the endothelium and suppressing the immune response, which in turn promotes the formation of mast cells and the deposition of sub-endothelial lipids [29]. We found MLR, NLR and SII higher in CAD patients in comparison with without CAD too (Table 11). Accordingly, SIRI was higher, and LMR – lower in our CAD patients’ group. It should be noted that Si Y. et al. also excluded patients with acute coronary syndrome, renal failure, connective tissue disease, severe valvular heart disease, and hypertrophic cardiomyopathy, but included patients with diabetes mellitus and ischemic stroke, and that these comorbidities may have had an impact on the calculated and tested inflammatory and total blood count readings [29].

In conclusion, we can state that LMR, NLR, MLR, MHR, SIRI and SII are available and biomarkers that could help assess the severity of coronary atherosclerosis in patients with SAP. Our study showed that NLR, SII, LMR, MLR and SIRI have strongest correlations with CAD severity. However, further research works is needed to determine the clinical relevance of SIRI, LMR, MLR and others in the evaluation of chronic inflammation in CAD, to confirm its usefulness in clinical practice, to establish standard cut-off values, and to elucidate the underlying mechanisms linking the readings to the pathophysiology of CAD. According to our knowledge, we firs evaluated correlation between all total blood count calculated readings related to chronic inflammation and severity of CAD in the same patient’s population. It should be emphasised that our investigated patients have not comorbidities, so the readings were not affected by other pathologies. Our findings confirm that SIRI and LMR could reflect the low chronic inflammatory statement in SAP patients best of all other readings and they are most reated to the degree of atherosclerosis in SAP patients. SIRI seems could reflects the degree of atherosclerosis best of all other calculated total blood count readings.

## Limitations

This study has several limitations. It is a pilot retrospective study evaluating the calculated total blood count readings related to low chronic inflammation as related with CAD severity in one small centre. Therefore, the results of the association between the severity of coronary artery stenosis and the readings level should be further tested in studies with larger sample sizes. Another limitation is that plaque composition was not assessed in this study (no intravascular ultrasound or optical coherence tomography was used, nor was plaque composition assessed by CCTA), as the type of plaque may also influence calculated total blood inflammation-related readings. Coronary artery calcium score was also not assessed. Finally, this was a short-term retrospective pilot study, so future adverse major cardiac events and mortality were not assessed.

## Conclusions

NLR, MLR, SIRI ans SII were higher and LMR was lower in patients with CAD. LMR decreased and NLR, MLR, SIRI and SII increased gradually with increasing in Gensini group number. LMR decreased and SIRI with MHR increased gradually with increasing in CAD-RADS group number. So, NLR, LMR and SIRI seems to be the best for chronic inflammation evaluation in patients with SAP.

It was found moderate correlation between inflammatory indexes SII, NLR and SIRI and severity of coronary stenosis according to Gensini score. The CAD-RADS score moderately correlated with LMR, MLR and SIRI. NLR, MLR and SIRI correlated with some exocardiography left ventricle parameters. So, SIRI seems could reflects the degree of atherosclerosis best of all. Easily measurable, and its low-cost ability to integrate multiple immune cells could makes it a valuable marker for assessing systemic inflammation, atherosclerosis progression and degree in SAP patients.

## Data Availability

All data produced in the present work are contained in the manuscript

## Author contributions

M.A. conceptualized the article; L.A. synthesized data; L.A. drafted the article; L.A. made the tables and figures; M.A. and L.J. contributed andcritically edited the article; all authors discussed the results and commented on the article.

## Acknowledgements.

This research received no external funding.

